# Efficacy, public health impact and optimal use of the Takeda dengue vaccine

**DOI:** 10.1101/2024.08.10.24311393

**Authors:** Bethan Cracknell Daniels, Neil Ferguson, Ilaria Dorigatti

## Abstract

Dengue is the most common arboviral infection, causing substantial morbidity and mortality globally. The licensing of Qdenga, a second-generation vaccine developed by Takeda Pharmaceuticals, is therefore timely, but the potential public health impact of vaccination across transmission settings needs to be evaluated. To address this, we characterised Qdenga’s efficacy profile using mathematical models calibrated to published clinical trial data and estimated the public health impact of routine vaccine use. We find that efficacy depends on the infecting serotype, serological status, and age. We estimate that vaccination of children aged over six years in moderate to high dengue transmission settings (seroprevalence at 9 years of age > 60%) could reduce the burden of hospitalised dengue by 10-22% on average over ten years. We find some evidence of a risk of vaccine-induced disease enhancement in seronegative vaccine recipients for dengue serotypes 3 and 4, especially for children under six years of age. Because of this, the benefits of vaccination in lower transmission settings are more uncertain, and more data on the long-term efficacy of Qdenga against serotypes 3 and 4 are needed.

## Introduction

With more than half of the world’s population currently at risk of dengue infection, novel control methods, including vaccines, are urgently needed to reduce disease burden and resulting economic impacts. Historically, developing safe and effective dengue vaccines has been challenging due to the presence of four antigenically distinct dengue serotypes (DENV1-4) that elicit cross-reactive immunity and can enhance the severity of secondary infections through antibody-dependent enhancement^1^. The first licensed dengue vaccine, Dengvaxia, developed by Sanofi Pasteur, was belatedly found to increase the risk of hospitalisation in dengue naïve (seronegative) vaccine recipients (hazard ratio: 1.75, 95% confidence interval (CI): 1.14 to 2.70)^2^, confirming earlier modelling of the phase III trial of that vaccine which had highlighted this potential risk^3^. Consequently, Dengvaxia is now only indicated for use in individuals with prior dengue exposure (seropositives), and due to the absence of an accurate rapid antibody test for dengue to date, is in limited use. There therefore remains an unmet need for a safe and efficacious dengue vaccine that can be used programmatically without pre-vaccination testing.

Previous work has shown that high neutralising antibody titres against dengue are associated with protection, and low-to-moderate antibody titres are associated with an increased risk of severe disease and hospitalisation ^4
–6^. However, an exact titre for protection has not yet been identified, and it is expected that this will depend on the assay used, the infecting serotype (and potentially genotype), and most likely an individual’s prior exposure to other serotypes and related flaviviruses^7,8^. Nevertheless, vaccine-induced antibody titres correlate with protection at the population level^9^ and are good predictors of disease risk^10^, leading to the World Health Organisation (WHO) recommending the use of neutralising antibody titres as immunogenicity metric for second-generation dengue vaccines^11^.

Qdenga, a second-generation vaccine developed by Takeda Pharmaceuticals, has recently been approved for use in several countries, including Brazil where vaccine rollout began in early 2024^12^. Qdenga is a tetravalent chimeric live-attenuated vaccine using DENV2 as the backbone for all four serotype components, but substituting DENV1, 2 and 4 pre-membrane and envelope proteins for those serotypes^13^. Qdenga’s efficacy was evaluated in a multi-country phase III trial across Asia and South America that enrolled approximately 21,000 participants aged 4-16 years, who were randomised 2:1 to receive two doses of Qdenga or placebo, 90 days apart^14^. Building on the experience with Dengvaxia, vaccine efficacy (VE) was evaluated by baseline serostatus prior to vaccination, infecting serotype, age, and disease outcome (defined as symptomatic dengue and hospitalisation) for all trial participants, at 12 ^14^, 18 ^15^, 24 ^16^, 36 ^17^, and 54 months ^18^ post-second dose.

From 1-57 months post-first dose, the average VE in the safety population (individuals who received at least one dose of the vaccine or placebo) was estimated at 61.2% (95% CI: 56.0 to 65.8) against symptomatic virologically confirmed dengue (VCD) and 84.1% (95% CI: 77.8 to 88.6) against hospitalised VCD over all serotypes and baseline serostatuses^18^. However, VE waned over time, from an average of 80.2% (95% CI: 73.3 to 85.3) against symptomatic VCD in the per-protocol population (individuals without any major protocol violations, including not receiving both doses of the correct assignment of Qdenga or placebo) in year 1 to 44.7% (95% CI: 32.5 to 54.7) in year 3. VE also varied by serotype and baseline serostatus, with higher VE in seropositive individuals and against DENV2^17^. Consistent with the VE estimates, neutralising antibody titres induced by Qdenga were highest and more durable in seropositive individuals and against DENV2^18^ with specific antibody^19^ and T cell^20,21^ responses most strongly elicited against the DENV2 backbone virus.

In baseline seronegative individuals, the phase III trial showed no statistically significant evidence of protection against DENV3 and DENV4, with average VE estimates up to 57 months post-first dose of 15.5% (95% CI: -108.2 to 35.9) and -105.6% (95% CI: -628.7 to 42.0), respectively^18^. During this period, point estimates of VE against hospitalisation following a DENV3 infection were negative in seronegative recipients (VE: -87.9%, 95% CI: -573.4 to 47.6), although 6 out of the 11 DENV3 hospitalisations in seronegative vaccinees occurred in Sri Lanka, where hospitalisation tends to be more frequent than in other countries^18^. Additionally, there were two cases of severe dengue in the seronegative vaccine group, both DENV3, and none in the placebo group^22^. Using the WHO 1997 criteria for dengue haemorrhagic fever, there were two cases in the seronegative placebo group (DENV1 and DENV3) and four in the vaccine group (all DENV3). This raises the question of whether these uncertain estimates represent a weak signal of a risk of vaccine-associated disease enhancement in seronegative recipients for DENV3 and DENV4, despite the lack of statistical significance.

While the published VE estimates for Qdenga provide estimates of how VE vary by serotype, age, serostatus and over time, most such estimates are only available stratified by at most two of those variables. For instance, no efficacy estimates have been published by both serotype and age to date. To support optimal deployment, it is also important to estimate the public health impact of Qdenga vaccination and evaluate its suitability across different transmission settings. Here we address these knowledge gaps. We first develop a survival model calibrated to published phase III data to infer how antibody titre dynamics can be translated into estimates of protection. Second, we embed this VE model into a previously published dengue transmission model^3^ to simulate the potential public health impact of programmatic use of Qdenga, estimating impacts at both the population-and individual-level. This work informed the WHO Scientific Advisory Board of Experts (SAGE) recommendations on dengue vaccines^23^ and the latest WHO position paper on dengue vaccination^22^.

## Results

### Vaccine efficacy against symptomatic disease and hospitalisation

We fitted a Bayesian cohort survival model to all published Qdenga phase III clinical trial data including symptomatic and hospitalised cases (**Supplementary Figures 1a and 2**). The model extends the correlates of protection model first proposed by Khoury *et al*.^24^ for SARS-CoV-2, allowing us to link the neutralising antibody titres induced by Qdenga (**Supplementary Figure 3**) to the risk of disease in the vaccine arm compared to the placebo group. Of the 31 model variants explored (**Supplementary Figure 4**), the optimal model following model selection reproduced the symptomatic case and hospitalisation attack rates observed over the 54 months post-second dose by inferring the relative serotype-, serostatus-, outcome-and age-specific titres required for protection and testing the hypothesis of potential vaccine-associated enhancement in seronegative individuals. We fit the model to the reported number of cases stratified by trial arm, age group, serotype, and baseline serostatus at the finest granularity allowed by the published data (**Figure 1a-c, Supplementary Figures 5-7**). Posterior parameter estimates are given in **Supplementary Table 1**.

**Figure 1:**
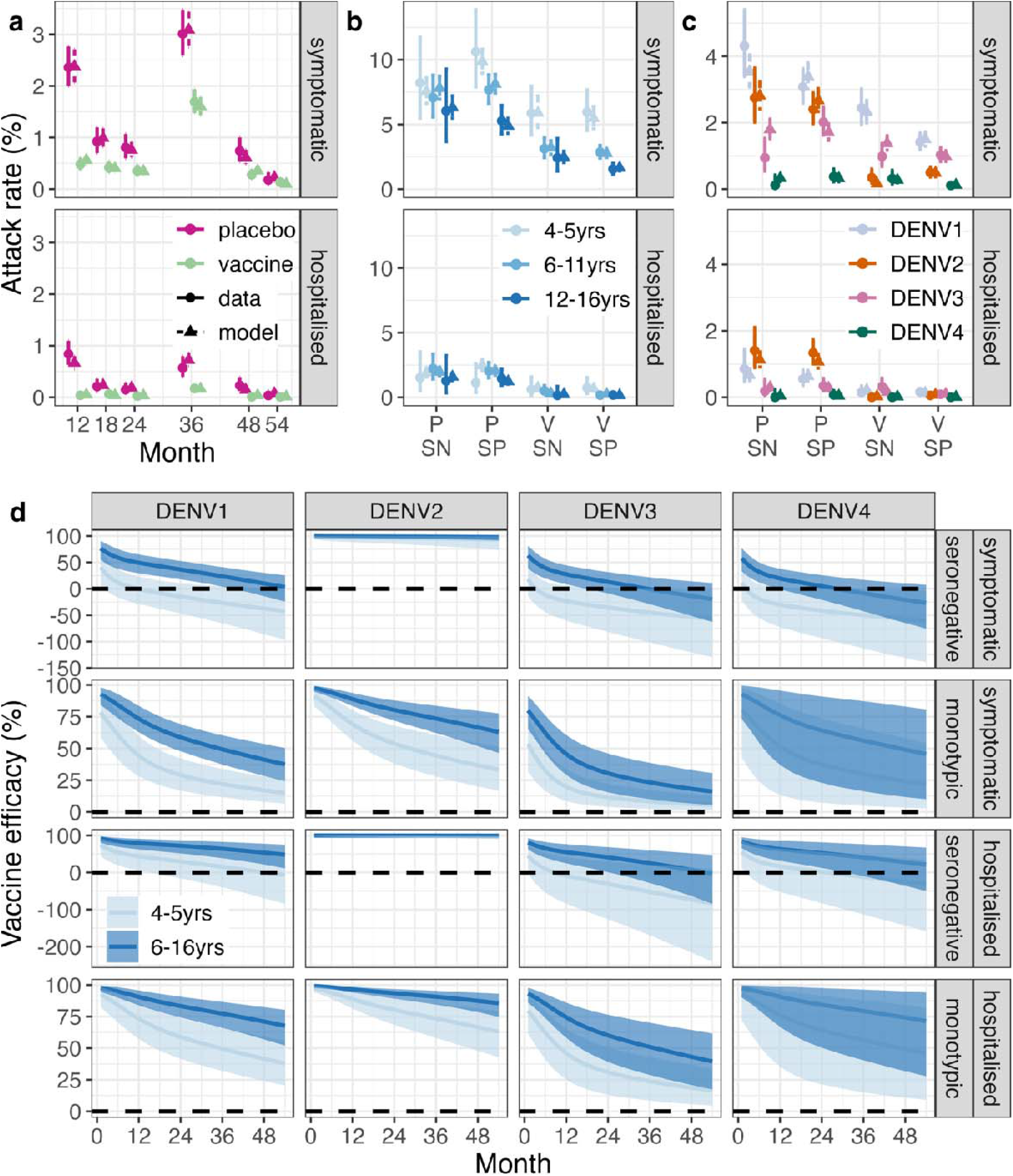
Attack rate and vaccine efficacy estimates. Observed and estimated attack rates of symptomatic virologically confirmed disease and hospitalisations in the phase III clinical trial by (**a**) trial arm and time, (**b**) age, serostatus, and trial arm and (**c**) serotype, serostatus, and trial arm. Note that the plotted resolution is lower than the data used in the model calibration. The modelled attack rates show the mean (triangle) and 95% Credible Interval (CrI, dashed line). The observed attack rates show the mean (circle) and 95% exact binomial confidence interval (solid line). (**d**) Estimated vaccine efficacy by serotype (columns), serostatus (rows), age group (colours), against symptomatic disease and hospitalisation (rows). The solid line represents the mean of the posterior distribution and the shaded area represents the 95% CrI. The dashed horizontal line marks 0 efficacy. VE in multitypic individuals is shown in Supplementary Figure 8. SN: seronegative. SP: seropositive. V: vaccine arm. P: placebo arm.

We estimate high VE against symptomatic and hospitalised DENV2 in both seronegative and seropositive individuals, regardless of age (**Figure 1d** and **Supplementary Figure 8**). In seropositive individuals, we estimate more moderate protection against symptomatic disease and hospitalisation for the other serotypes. Conversely, for seronegative vaccinated subjects in the 4-5 years age group, we estimate an enhanced risk of symptomatic disease (negative average VE) following DENV1, DENV3, and DENV4 infection, starting from 3-12 months post second dose, albeit credible intervals always include zero. For DENV3 and DENV4 we also estimate an enhanced risk of hospitalisation from 9- and 26-months post second dose in the seronegative 4-5-year age group, respectively. For seronegative children over 6 years old, we estimate a positive average VE against both symptomatic disease and hospitalisation for DENV1 for the entire follow up period (54 months post second dose) but negative average VEs were estimated for DENV3 and DENV4 from approximately 3 years post second dose. The large uncertainties around the VE estimates against hospitalisation and for DENV3 and DENV4 are due to the small case numbers observed during the trial. The VE estimates projected up to 15 years post-vaccination as a function of time and neutralising antibody titre are shown in **Supplementary Figures 9 and 10**.

Our model explicitly included a parameter determining the maximum potential level of vaccine-associated disease enhancement, which was estimated to be 84% (95% CrI: 13% to 184%). The Bayes factor comparing our model with and without disease enhancement indicates evidence for enhancement (**Supplementary Figure 11**) regardless of the prior distribution used, even though the magnitude of the estimated enhancement parameter was influenced by the choice of the prior distribution. The results of sensitivity analysis on the period of cross-protective immunity between serotypes and whether there is clinical disease in post-secondary infections are shown in **Supplementary Figure 12**. When fitting to simulated data, all parameters were recovered well (**Supplementary Figure 13**).

### Population impact of routine Qdenga vaccination

To estimate the impact of Qdenga vaccination, we integrated our fitted VE model (**Figure 1d**) into the multi-strain stochastic compartmental model of dengue transmission previously used to investigate the potential impact of Dengvaxia^3^. We explored four different hypotheses of the vaccine′s mode of action, combining two assumptions regarding protection (against disease only, VS, or also against infection, VI [**Figure 1D**]), with two assumptions about the duration of VE decay (up to 5 years, D5, or 15 years, D15) (**Supplementary Figures 9**). Given the limited data available^25^, we assumed that protection against infection requires a higher titre compared to protection against symptomatic disease and that vaccine-associated enhancement applies only to clinical outcomes^5^ (See **SI Section 3.3** and **Supplementary Figure 14**). We examine a range of transmission settings with different forces of infection (characterised by the average seropositivity of 9-year-olds, in line with our previous work^3^). For each transmission intensity level, vaccine mode of action, coverage level (20%, 40%, 60%, 80%), age of vaccination (4-12 years), and population demography (Brazil and the Philippines), we sampled 200 posterior parameter estimates from our VE model (**Figure 1d**) and for each such posterior sample we ran 50 simulations of the dengue transmission model, giving 10,000 simulations in total per scenario (**Supplementary Figure 1b**).

**Figure 2a** shows the population impact, summarised as the total proportion of cases averted over 10 years, assuming the Brazilian demography, 80% vaccination coverage in 6-year-olds, and that VE wanes for 15 years (scenarios VS_D15 and VI_D15). **Supplementary Figures 15-17** show that the expected impacts assuming the Philippines demography and that antibody waning lasts for 5 years, are similar to the scenario presented in the main analysis (**Figure 2a**). Under the VS assumption, we find that the population-level impact increases as the intensity of transmission increases whilst under the VI assumption the population-level impact is similar across all transmission settings with <50% seropositive 9-year-olds on average (**Figure 2a**). Regardless of the transmission setting or VE against infection, the population impact is modest, with the mean proportion of symptomatic cases prevented over 10 years ranging from 1.6% to 13.7% under the VS scenario and from 8.9% to 17.2% under the VI scenario, depending on transmission intensity. The mean proportion of hospitalisations averted is slightly higher, rising to 22.4% (95% CrI: 17.8% to 28.3%) assuming the VI scenario in the highest transmission settings.

**Figure 2:**
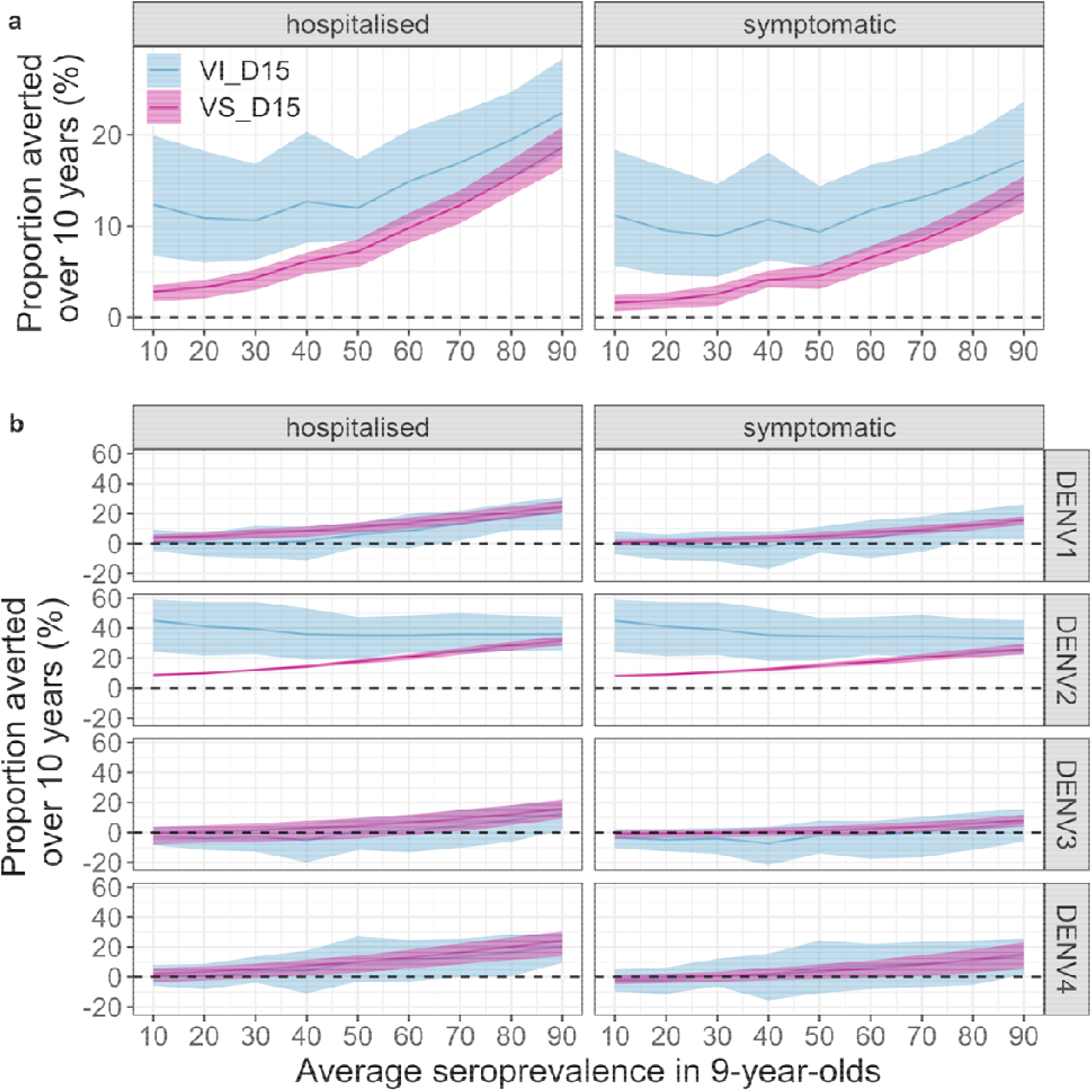
Population-level impact of vaccination. Cumulative proportion of hospitalised and symptomatic cases averted over 10 years (y-axis) by transmission setting (x-axis, expressed as the average seroprevalence in 9-year-olds), assuming efficacy against infection and disease (VI, blue) or only against disease (VS, pink) decaying for 15 years (D15), using 80% coverage across ten years and the Brazilian demography (**a**) over all serotypes and (**b**) by serotype. The solid line represents the mean, the shaded regions represent the 95% credible interval.

**Figure 2b** shows the population impact by serotype, which highlights the potential for small negative impacts against DENV1, DENV3, and DENV4 in low-to-moderate transmission settings. In other words, in low-to-moderate transmission settings, most of the positive impact of vaccination observed at the population level in both the VI and VS scenarios can be attributed to the prevention of DENV2 cases (**Figure 2b**).

**Figure 3** and **Supplementary Figure 18** present the population impact by age of vaccination in the Philippines and Brazil, respectively. In low to moderate transmission settings the impact is relatively insensitive to the age of vaccination, especially under the VS assumption (**Figure 3**). In moderate to high transmission settings, the optimal age of vaccination decreases from age 11 to 6 as average 9-year-old seropositivity increases from 50% to 90%, under the VS assumption (**Figure 3**). Under the VS assumption, reductions in the coverage from 80% to 20% reduce impact proportionately (i.e., by 75%) regardless of the transmission setting, whereas under the VI assumption, reducing coverage from 80% to 20% reduces the impact by between 68% to 74%, depending on the transmission intensity (**Supplementary Figure 19-20**).

**Figure 3:**
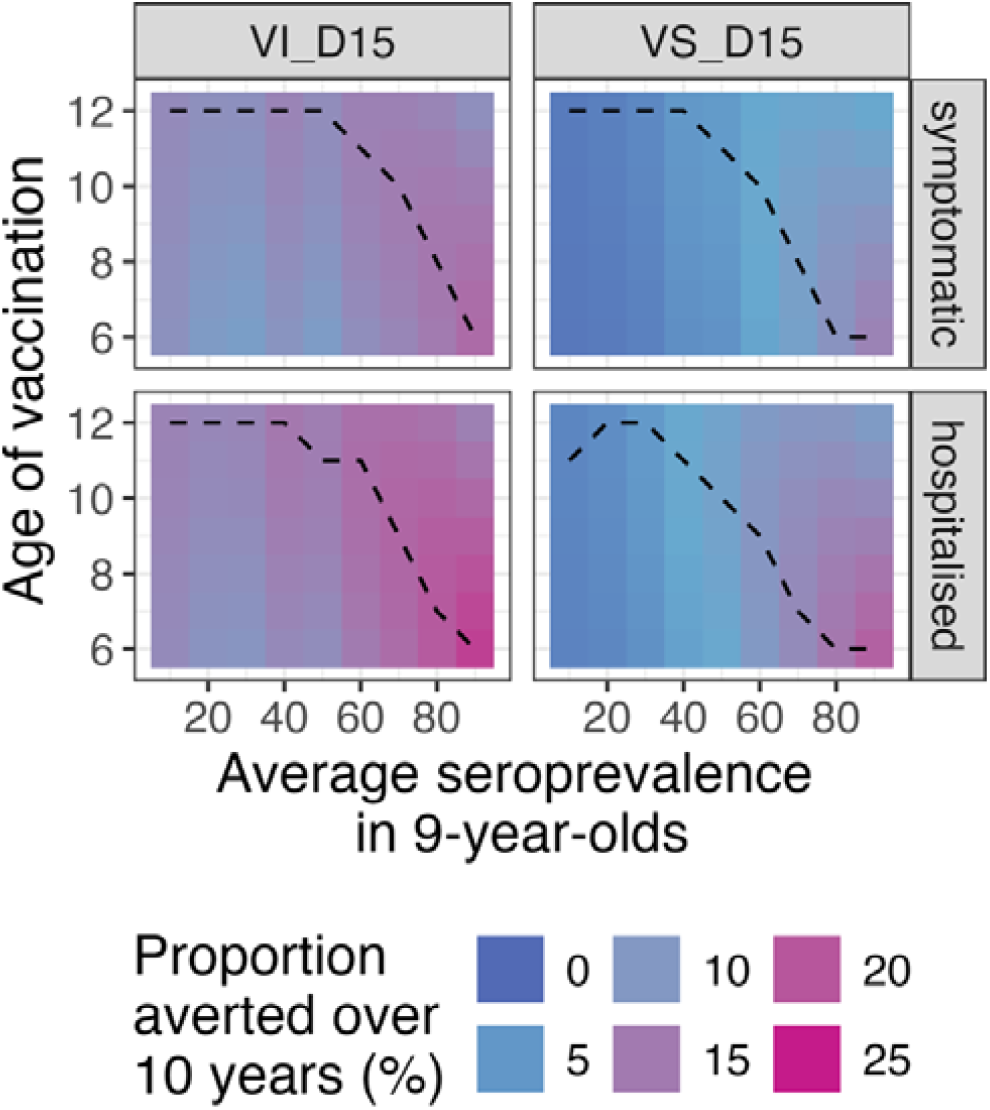
Impact of age at vaccination on the population-level impact by transmission setting. Cumulative proportion of hospitalised and symptomatic cases averted (rows) by transmission setting (x-axis, expressed as the average seroprevalence at 9-year-old), and vaccine mechanism (columns) assuming vaccination of ages 6-12 (y-axis) and the Brazilian demography, over ten years. VI_D15: scenario assuming efficacy against infection and disease decaying for 15 years post vaccination. VS_D15: scenario assuming efficacy against disease decaying for 15 years post vaccination.

### Individual-level impact of routine Qdenga vaccination

Whilst the overall average population impact is always positive, the individual benefits and risks of vaccination - measured as the proportion of cases averted in the first vaccinated cohort over ten years - show a more complex picture (**Figure 4** and **Supplementary Figures 21-23**). The mean individual impact is positive, with 20-47% and 42-68% of symptomatic cases and hospitalisations averted in vaccinated individuals over 10 years, respectively. Seropositive individuals always benefit from vaccination, with 40-55% and 63-75% of symptomatic and hospitalised cases averted, respectively. Conversely, whilst the mean impact is positive for seronegative vaccinees, negative impacts in low-to-moderate transmission settings are possible, as demonstrated by the negative lower bound of the 95% uncertainty intervals (**Figure 4**) reflecting both the uncertainty in the VE estimates (**Figure 1d**) and in the circulating serotypes across the simulations.

**Figure 4:**
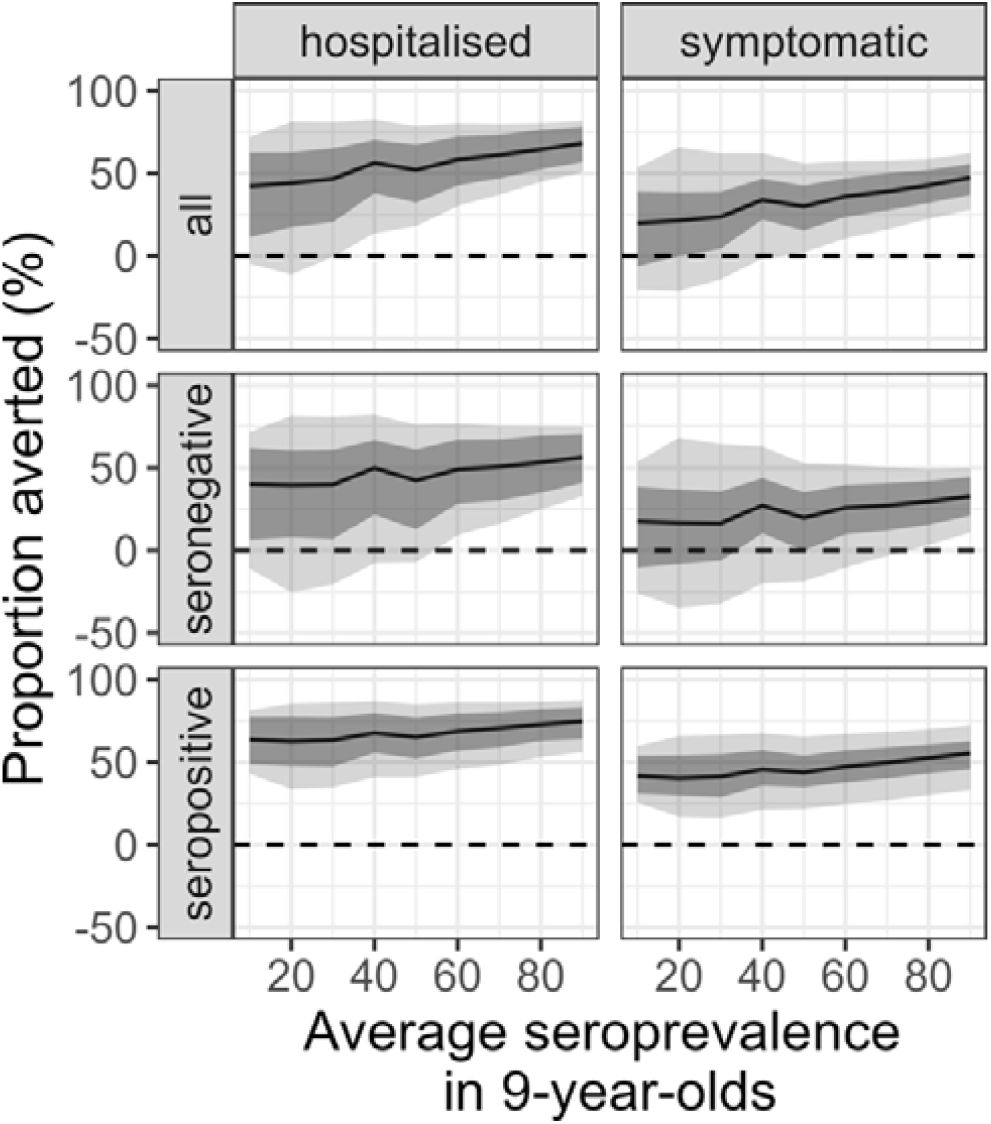
Individual-level impact of vaccination. Proportion of hospitalised and symptomatic cases averted (columns) in the first vaccinated cohort of 6-year-olds over ten years by transmission setting, expressed as the average seroprevalence at 9-year-old (x-axis) assuming a vaccination coverage of 80% using model VS_D15 (efficacy against disease decaying for 15 years post vaccination) and the Brazilian demography. The impact is shown overall (all) and among baseline seropositive and seronegative vaccinees (rows). The solid lines represent the mean, light shading represents the overall uncertainty (95% CrI), and the dark shading represents the parameter uncertainty (95% CrI).

**Figure 5a** shows that overall, vaccination is expected to avert 95 (95% CrI: 25 to 178) cases and 14 (95% CrI: 1-23) hospitalisations (assuming 9% of symptomatic cases are hospitalised) per 1,000 vaccinated individuals in moderate transmission settings (at least 60% seropositive 9-year-olds on average), estimates which are driven primarily by the prevention of cases in seropositive individuals. In seronegative individuals, symptomatic cases are averted almost entirely against DENV2 (**Figure 5b**), regardless of the transmission setting. Notably, negative DENV3 impacts are more likely than positive in seronegative individuals in transmission settings with <60% seropositive 9-year-olds on average.

**Figure 5:**
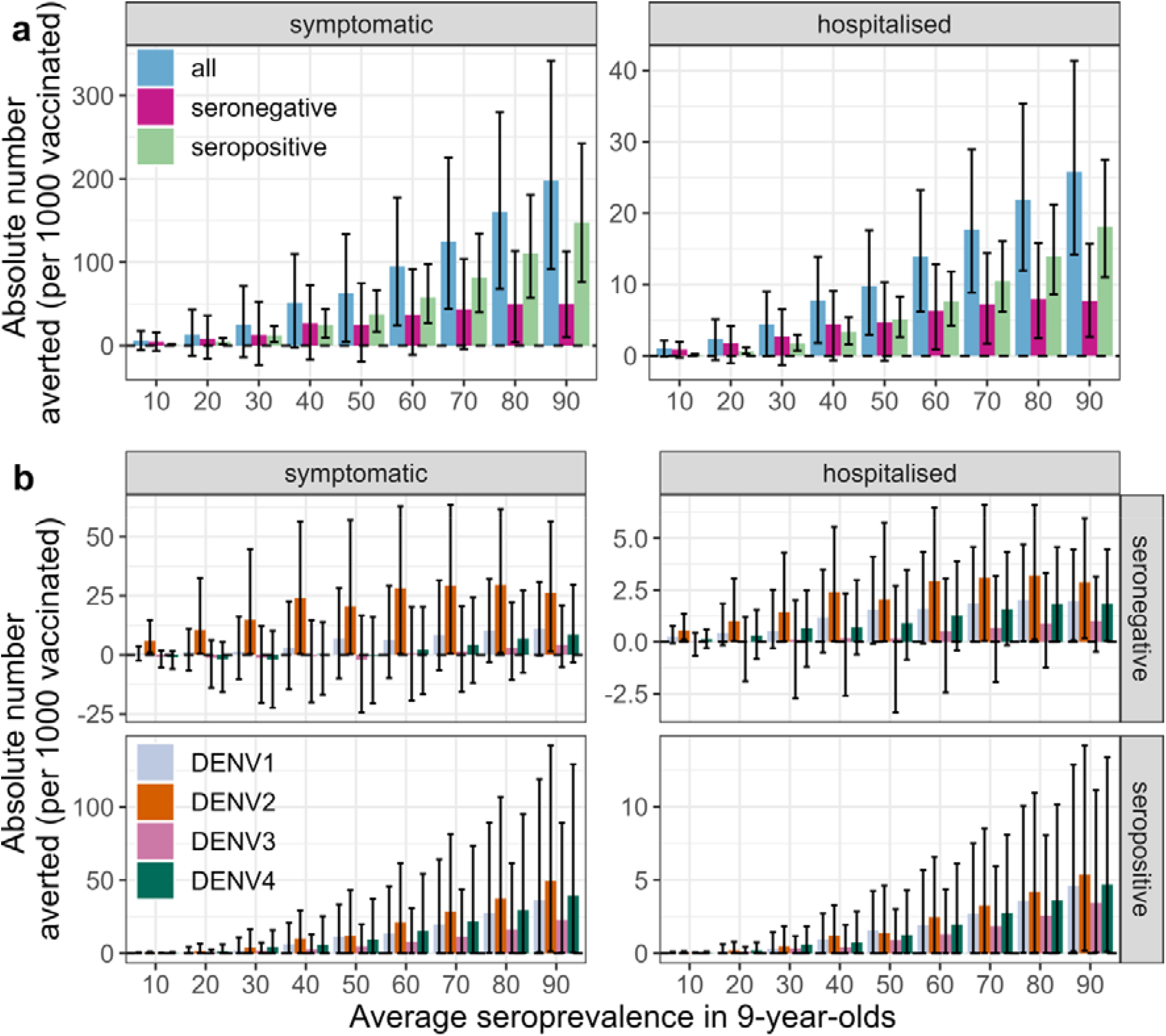
Cases averted by vaccination per 1,000 vaccinated children. Absolute number of symptomatic cases and hospitalisations (columns) averted over 10 years post second dose (y-axis) in the first vaccinated cohort of 6-year-olds per 1,000 fully vaccinated persons by transmission setting, expressed as the average seroprevalence in 9-year-olds (x-axis), assuming model VS_D15 (efficacy against disease decaying for 15 years post vaccination), the Brazilian demography and that 9% of symptomatic cases are hospitalised. (**a**) Cases averted in the entire vaccinated cohort (all, blue), and among baseline seropositive (green) and baseline seronegative (pink) vaccinees and (**b**) cases averted by serotype (colours) and serostatus (rows). The bars represent the mean, and the error bars represent the overall uncertainty (95% CrI).

### Population screening

A potential strategy to mitigate the risk in seronegative recipients (**Figure 4**) is through pre-vaccination serological screening, as recommended for Dengvaxia^11^. Assuming a diagnostic test with 94.7% specificity and 89.6% sensitivity^26^, and either the VI or VS mode of vaccine action, we find that pre-vaccination screening reduces the population impact of vaccination on symptomatic disease and hospitalisations by 29-33% in the highest transmission setting and 82-85% in the lowest transmission setting (**Supplementary Figure 24-25**). As a result, the proportions of symptomatic and hospitalised cases averted over 10 years are <15% across all transmission settings when implementing pre-vaccination screening. This driven by the loss of protection against DENV1 and DENV2 in seronegative recipients (**Figure 5b**). The impact of pre-vaccination screening on vaccinated individuals is shown in **Supplementary Figures 26-27**. Overall, screening increases the individual impact, as a higher proportion of the vaccinated individuals are seropositive. In the seronegative individuals who are vaccinated due to imperfect test specificity, screening does not change the proportion of cases averted.

### Impact of vaccination on serotype dynamics

Given the imbalanced serotype-specific efficacy profile (**Figure 1d**) and population impacts (**Figure 2b**), we investigated the extent to which vaccination assuming the VI scenario (where vaccination affects transmission) could impact serotype-specific dengue dynamics. Specifically, we tested whether the introduction of the Qdenga vaccine could (i) eliminate DENV2 in settings with low DENV2 circulation and favour the circulation of the other serotypes and (ii) increase the dominance of DENV3 in settings with high DENV3 circulation. We investigated changes in the serotype-specific dynamics among the 1,000 simulations that, in the absence of vaccination, had the lowest and highest DENV2 and DENV3 incidences, respectively. **Figures 6a-b** show example transmission dynamics with and without vaccination and demonstrate how, under the VI assumption, outbreak peaks are expected to change in both timing and magnitude, especially in settings with higher transmission intensities.

**Figure 6:**
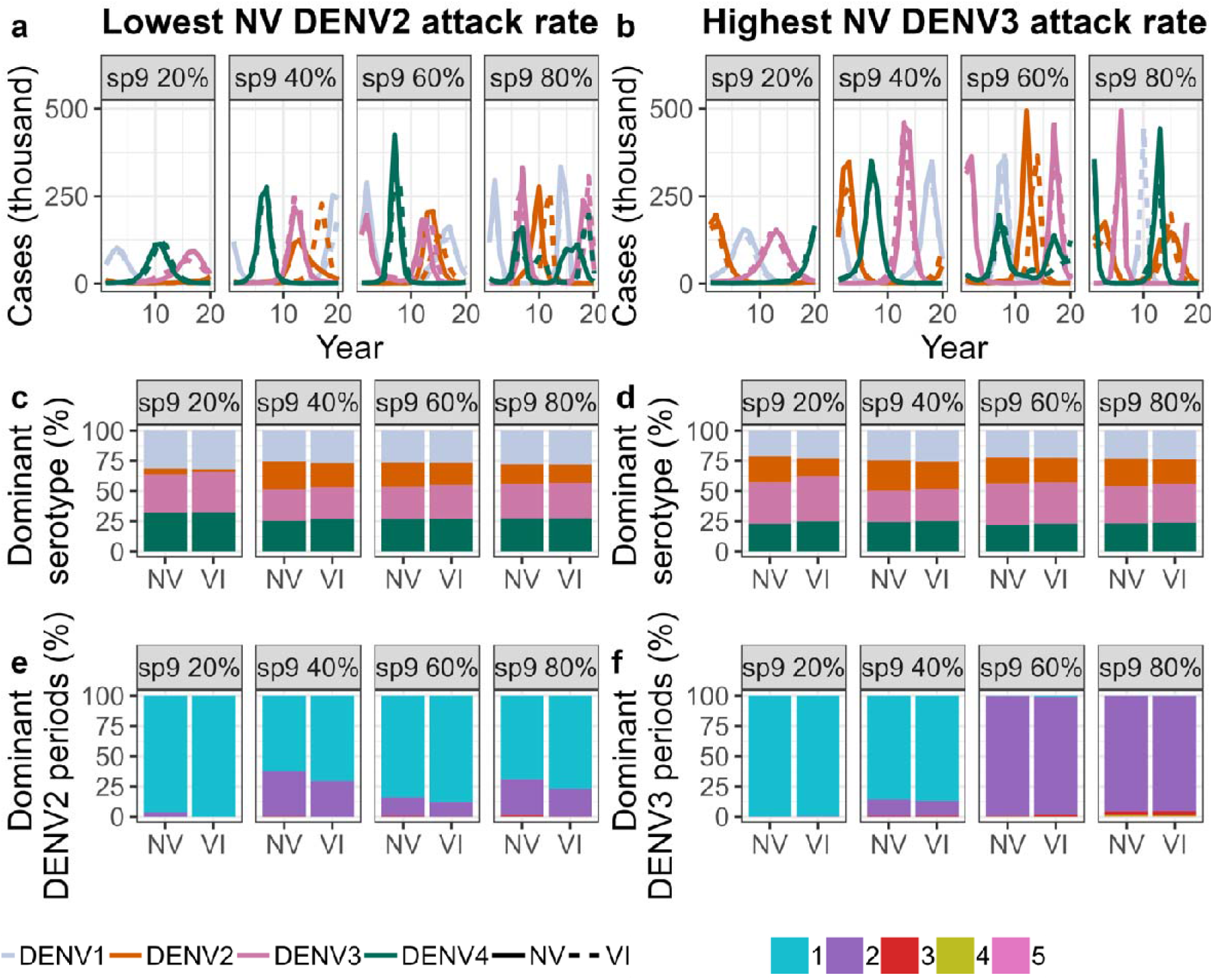
Impact of vaccination on serotype dynamics. For the 1,000 simulations with the lowest DENV2 burden **(a, c, e)** and the highest DENV3 burden **(b, d, f)**, across transmission settings (sp9, average seroprevalence in 9-year-old) with (VI) and without (NV) vaccination we show **(a-b)** example serotype-specific transmission dynamics **(c-d)** the mean proportion of time each serotype is dominant and **(e-f)** the proportion of simulations (y-axis) in which DENV2 **(e)** and DENV3 **(f)** become the dominant serotype for the specified number of seasons (colours), over a 20 year period since the start of vaccination.

In settings with low-to-moderate transmission intensity with limited DENV2 circulation, we expect vaccination to reduce slightly DENV2 dominance, with a lesser impact in higher transmission settings (**Figure 6c-e**). Conversely, we estimate that in low transmission settings with high DENV3 circulation, the introduction of Qdenga may increase DENV3 dominance, but in high transmission settings this effect is modest (**Figure 6d**) and the number of DENV3 dominant periods over 20 years remains largely unchanged by vaccination (**Figure 6f**). Taken together, these results suggest that under the VI mode of vaccine action, routine vaccination would only minimally alter the serotype-specific dynamics that would be observed in the absence of vaccination.

## Discussion

By combining a correlates-of-protection model of VE with a mechanistic model of dengue transmission, we show that Qdenga has an efficacy profile and predicted public health impact that varies with age, serostatus, and infecting serotype. We estimate that Qdenga is highly protective against DENV2 regardless of serostatus and moderately protective against the other three serotypes in seropositive individuals, with evidence of potential enhancement in disease risk following DENV3 and DENV4 infection in seronegative subjects. Notably, we found that this risk is greatest in seronegative children aged 4-5 years at vaccination. Despite this imperfect efficacy profile, our estimates suggest that the introduction of Qdenga could reduce the burden of hospitalised dengue in high transmission intensity settings by up to approximately 20% over the first ten years of routine programmatic use. These findings support the recent WHO recommendations on the use of Qdenga in children aged 6 years of age and above in settings with high dengue disease burden and transmission^22,27^.

Qdenga VE estimates stratified by serotype and age have not yet been reported by the phase III trial investigators^14–18^. However, our model-derived estimates agree with early clinical studies and previous work on the functional immune response elicited by Qdenga which suggested that DENV2 is the dominant replicating serotype and driver of homotypic immunity^19,20,28^. We find that Qdenga protects against symptomatic disease and hospitalisation caused by DENV1, DENV3 and DENV4 in seronegatives aged 4-5 years for 3-15 months, after which we estimate a potential risk of disease enhancement. In seronegative children aged 6-16 years, we estimate the initial period of partial protection to be at least 27 months. The uncertainty around VE and potential risks for seronegative recipients highlight the need for continued monitoring of Qdenga’s efficacy profile and collection of additional data in post-licensure studies^23,29^, especially for DENV3 and DENV4. The use and timing of an additional booster dose is also currently being evaluated^22^ and it will be important to assess whether an extra vaccine dose might mitigate the potential risks that we have identified.

We cannot rule out a potential risk of enhancement in any seronegative subjects, but we found that VE was higher and the risk of enhancement lower in children over 6 years of age compared with those under 6, independent of their serostatus. Age-dependencies in VE were found also in the analysis of the Dengvaxia clinical trial data^30,31^ and whilst the reasons for this age-dependency are not yet fully understood, these results suggest reduced antibody and cell-mediated memory responses to vaccines in young children^32,33^.

Our analysis shows that population impact and individual benefit/risk depend on setting-specific transmission intensity and dominant circulating serotypes. In low transmission intensity settings, the average impact in seronegative individuals is positive but the positive impact is driven almost entirely by preventing DENV2 cases and is counterbalanced by lower (and potentially negative) impacts against DENV3 and DENV4. However, in higher transmission intensity settings (>60% seropositivity in 9-year-olds on average), the lower 95% credible bound on individual impact remains positive. Avoiding potentially negative individual impacts therefore requires Qdenga vaccination strategies tailored to local settings, using recent force-of-infection estimates^34^ and optimised vaccination ages. Pre-vaccination screening^35^ is expected to reduce the (already modest) population impact by about 30-80%, depending on the transmission setting and considering test limitations^36^, additional costs, logistical challenges, and the experience with Dengvaxia^37^, pre-vaccination screening for Qdenga is currently not recommended by WHO^22^.

In early 2024, Brazil became the first country to roll out Qdenga^12^ and our estimates suggest that, assuming 80% coverage, in a high transmission setting (60% seropositive 9-year-olds) Qdenga could avert 95 (95% Cr1: 25 to 178) symptomatic cases and 14 (95% Cr1: 1-23) hospitalised cases, per 1,000 children vaccinated. Careful monitoring of the Brazilian programme will be important to evaluate programme effectiveness, impact and safety.

Given the general lack of serotype-specific transmission intensity estimates, we assumed that all serotypes were approximately equally transmissible. However, during Qdenga’s phase III trial, spanning 4.5 years across eight countries in Southeast Asia and South America, the four serotypes (DENV1, DENV2, DENV3 and DENV4) accounted for 42.9%, 31.4%, 21.8%, and 3.9% of cases, respectively. If such patterns of serotype dominance continue in the future, the positive impact of Qdenga may be higher than estimated here. However, we note that DENV3 has recently re-emerged in several South American and Asian countries after its absence over several years, suggesting a need for caution in assuming that past serotype dominance trends will continue in the future. More generally, there is a need for better characterising the fundamental transmissibility and severity of the four serotypes. Critically, despite the estimated serotype imbalance in efficacy, our modelling suggests that even widespread programmatic use of Qdenga will at most cause minor changes in patterns of serotype dominance.

While modelling cannot substitute for the lack of data, our biologically motivated approach to VE modelling allowed us to share model parameters across strata, increasing statistical power. Although absolute dengue correlates of protection are yet to be identified, our analysis indicates that mean antibody titres kinetics can explain the efficacy of Qdenga, suggesting that neutralising titres are a surrogate marker for protection (or risk) from symptomatic and hospitalised dengue at the population level^5,6,10^. However, there are several limitations to our VE model. We needed to assume different threshold titres for protection for seronegative and seropositive vaccine recipients, suggesting that qualitative differences in the humoral response and potentially other immune functions, such as cellular immunity, can play an important role^19,20,38^. The lack of individual-level data meant that we could not investigate the link between individual antibody trajectories and vaccine induced protection, nor could we investigate how boosting of antibody titres due to subclinical infections and homotypic dengue re-exposure during the phase III trial ^10^ affected our VE estimates. Furthermore, the use of hospitalisation as a clinical endpoint (combined with the limited country-level data on hospitalisation protocols) limited the extent to which we could account for different country-specific rates of severe dengue disease.

The VE model developed in our study can be easily adapted to estimate and compare the efficacy of other dengue vaccines, such as the live-attenuated Butantan-DV vaccine currently being evaluated in Brazil^39^. The transmission model presented in this study can similarly be used to assess the potential impact of other dengue interventions and combinations of interventions, including new dengue vaccines, Wolbachia^40^, and antivirals ^41^, supporting the WHO recommendation to consider dengue vaccination as part of an integrated strategy to control dengue^22^. It will be important to evaluate how combinations of interventions will affect transmission dynamics, and hence the impact of vaccination.

In conclusion, this study finds evidence of high efficacy of Qdenga vaccination against DENV2 and against the other serotypes in seropositive individuals, resulting in modest reductions in dengue cases and hospitalisations across different transmission intensity settings. Conversely, except for DENV2, we found evidence for a potential risk of enhancement in seronegative individuals, especially in those aged 4-5 years at vaccination. The analysis presented in this paper informed the recommendations recently published in the WHO position paper^22^ on dengue vaccination and demonstrates how modelling can help translate clinical trial data and complex efficacy profiles into population impact estimates, thereby optimising vaccination deployment to minimise individual risk and maximise public health benefit.

## Data availability

All data used in the modelling study are on GitHub at https://github.com/bnc19/qdenga_impact.

## Code availability

Code is available at: https://github.com/bnc19/qdenga_impact.

## Acknowledgments

ID acknowledges funding from Wellcome Trust (213494/Z/18/Z). NMF, BCD and ID acknowledge funding from the MRC Centre for Global Infectious Disease Analysis (reference MR/X020258/1), funded by the UK Medical Research Council (MRC). This UK funded award is carried out in the frame of the Global Health EDCTP3 Joint Undertaking. NMF also acknowledges funding by the National Institute for Health and Care Research (NIHR) Health Protection Research Unit in Modelling and Health Economics, a partnership between the UK Health Security Agency, Imperial College London and LSHTM (grant code NIHR200908), and from the Jameel Institute provided by a philanthropic donation by Community Jameel.

Disclaimer: “*The views expressed are those of the author (s) and not necessarily those of the NIHR UK Health Security Agency or the Department of Health and Social Care*.”

## Online Methods

### Data

Published phase III clinical trial data are available for 12^14^, 18^15^, 24^16^, 36^17^, and 54 months^18^ post-30 days after the second dose endpoints. From these publications, we extracted the number of symptomatic VCD cases (*N*_*symp*_) and the number of VCD cases leading to hospitalisation (*N*_*hosp*_) at the finest stratification available. Case data were obtained by trial arm *v* (1 = placebo, 2 = vaccinated), and by baseline serostatus *b* (-= seronegative, + = seropositive), infecting serotype *k* (DENV1 to DENV4) and age group *j* (1 = 4-5yrs, 2= 6-11yrs, 3 = 12-16yrs), within each reporting interval *d* (1= 1-12 months, 2 = 13-18 months, 3 = 19-24 months, 4 = 25-36 months, 5 = 37-48 month, 6 = 49-54 months), as published. As only the first VCD case in each trial participant is reported in the phase III trial publication, we right-censored cases at the end of each reporting interval *d*. See **SI Section 1** for a complete overview of the data.

### Cohort survival model of VE

To estimate VE, we calibrated a Bayesian cohort survival model to the above data (**Supplementary Figure 1a**). We assumed an individual may be infected up to four times, and any infection could be symptomatic. We did not track further infections among individuals who had a detected infection. The model was solved at monthly time intervals (*t*= 1 to 54).

#### Fitting the initial conditions

Let *p*_*kj*_ denote the probability of exposure to serotype *k* (= 1,2,3,4) in age group *j* prior to the start of the trial (*T*_0_) and let *c* denote the number of serotypes an individual had been exposed to (= 0,1,2,3,4).

The probability that an individual has been exposed to serotypes Ω within seth_c_, defined as *h*_0_ — {Ø}, *h*_1_ = ({1}. {2}, {3}, {4}}, *h*_2_ = ({1,2}, (1,3}, {1.4}, {2,3}, {2,4}, {3,4}}, *h*_3_ = ({1,2,3}, {1,2,4}, {1,3,4}, {2,3,4}} and *h*_4_ = {{1,2,3,4}} is given by:

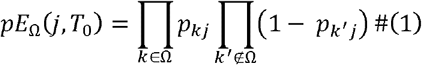

Accounting for the imperfect test sensitivity and specificity of the microneutralisation test used to classify individuals as seronegative (—) or seropositive (+) at baseline ^42^ (see **SI Sections 2.1 and 2.2** for full details), the probability of being classified as seropositive at baseline is:

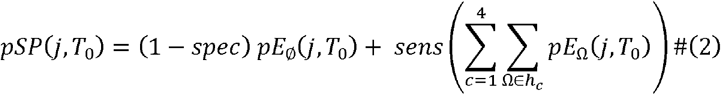

We estimate *pSP*(*j, T*_0_) by fitting the number of baseline seropositive individuals by age group *j* across both trial arms, *N*_*Sp*_*(j, T*_*0*_) to the observed data, assuming a Binomial distribution and the observed number of participants *N*_*pop*_ *(j, T*_*0*_) in age group *j* over both the vaccine and placebo arms:

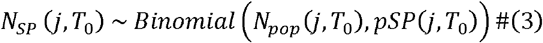

#### Estimating vaccine efficacy

We used a modified form of the correlates of protection model first proposed by Khoury et al. ^24^(to model VE for SARS-CoV-2 vaccines) to link the imputed neutralising antibody titre *n*_*ck*_(*t*)a gainst serotype *k* (**Supplementary Figure 3**) with the risk ratio (comparing vaccinated and unvaccinated individuals) of symptomatic disease 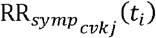 and hospitalisation 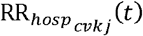:

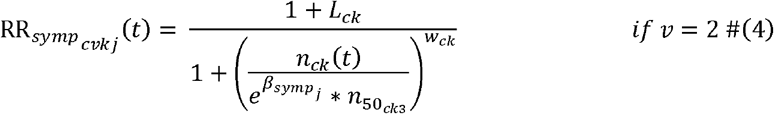

Eq. 4 presents the risk ratio of symptomatic disease, with the risk ratio of hospitalisation provided in the **SI Section 2.3**. By definition, 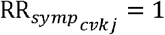 if *v* = 1. Here *L*_*ck*_ is the maximum potential vaccine-induced disease enhancement with serotype k associated with vaccination. This is assumed to be zero for all individuals with one or more previous DENV exposures (1 ≤ *c* ≤ 4), such that the maximum RR_*symp*_ is = 1 for all baseline seropositive vaccinees. We estimate *L*_*ck*_ (≥ 0) for vaccinated individuals with no prior DENV exposure (*c* = 0), such that their maximum RR_*syznp*_ may be >1. The parameter 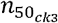 represents the neutralising titre conferring 50% protection against disease (in the absence of enhancement) for the oldest age group (*j* = 3), which is scaled by 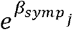 for individuals in the younger age groups, *j* ∈ {1,2}. Finally, *w*_*ck*_ controls the steepness of the relationship between neutralising titre and the risk ratio. Note that since RR_*symp*_ dependent on the cumulative number of DENV exposures at month t, not the number of exposures prior to vaccination, we are assuming that the order of vaccination and infection does not matter. So, for example, a baseline monotypic vaccinee escaping infection up to month t has the same disease risk as a baseline seronegative vaccinee with a single breakthrough infection prior to month *t*.

We assume that the vaccine induced neutralising antibody titres are characterised by an initial period of fast decay, with half-life *hs*_*b*_ (decay rate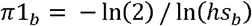), followed by a period of slow decay with half-life *hl* (decay rate 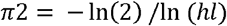

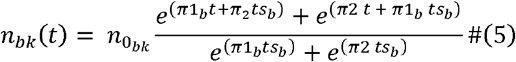

Here 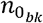 is the fitted initial neutralising antibody titre to serotype *k, b* is baseline serostatus prior to vaccination (*b* ∈ {—,+}), *ts*_*b*_ is the time at which decay switches from fast to slow, and t is the month elapsed since the second dose.

### Likelihood

Within each trial reporting period *d*, we assume a constant serotype-specific FOI over time, λ_*k*_(*d*). As the model is solved at monthly intervals t, we set λ_*k*_(*t*) = λ_*k*_(*d*) for all months t within the reporting period *d*.

For each month t, we calculate the probability of surviving that month without infection, given as 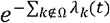 (**Supplementary Figure 1a**). The probability of being seronegative at the start of month *t* + 1 is therefore:

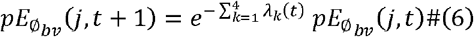

The probability of having been infected by the set of serotypes Ω by month *t* + 1 can be represented as the sum of those who were already exposed to the serotype combination in set Ω and who escaped infection by all serotypes not in 17 during month *t*, plus those with previous exposure to all serotypes in set Ω excluding serotype *k* (denoted Ω\_k_) who had an undetected infection to serotype *k* during month *t* — 12 (assuming a 12-month period of heterotypic cross-immunity and censoring individuals with detected infections following the date of detection^20,21^):

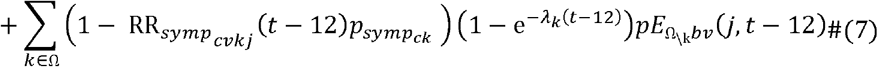

where the 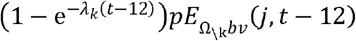 is the incidence of infection with serotype *k* during month *t*-12 in individuals previously exposed to all serotypes in set Ω excluding *k*. The probability of developing symptoms upon infection is denoted 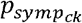. The second term in equation 7 represents the incidence of asymptomatic infection 12 months prior. Let r = the probability of symptoms during a secondary infection, ρ_*k*_ = the serotype-specific risk of symptoms during a primary infection relative to a secondary infection and *φ* = the risk of symptomatic disease during a post-secondary infection compared to a primary infection. Thus, 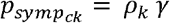 for primary infections (*c*= 0), = *γ* for secondary infections (*c* = 1), and= *ρ*_*k*_ *γ φ* for post-secondary infections (2 ≤ c ≤ 3).

The incidence of infection is:

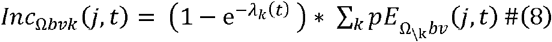

The incidence of disease and hospitalisation are:

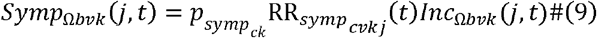

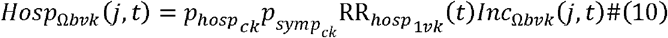

Here 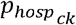 is the probability of hospitalisation. Let δ_*k*_ = the probability that a symptomatic case due to serotype k is hospitalised and ϵ = the risk of hospitalisation in secondary cases, compared to primary or post-secondary infections. Thus, 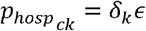 when *c*= 1, and *δ*_*k*_ otherwise. See SI Section 2.4 for additional details on the incidence calculations.

Summing over the serotype combinations, the total symptomatic case and hospitalisation incidence are:

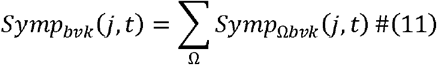

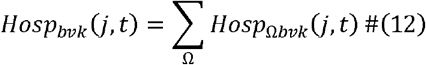

The monthly symptomatic and hospitalisation incidences are aggregated to match the months within each trial reporting period *d*, and the expected number of symptomatic and hospitalised cases are calculated as:

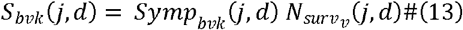

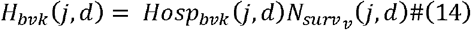

where 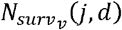 is the observed population size in reporting period *d*, accounting for censoring of symptomatic cases in the previous time intervals. The observed total number of symptomatic cases and hospitalisations at time interval d are assumed to follow Binomial distributions:

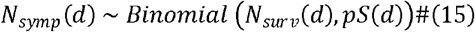

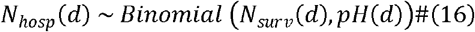

where *pS*(*d*) = ∑_*b*_ ∑_*v*_ ∑_*k*_ ∑_*j*_ S_*bvk*_ (*j,d*)/*N*_*surv*_ (*d*) and *pH*(*d*) = ∑_*b*_ ∑_*v*_ ∑_*k*_ ∑_*j*_ H_*bvk*_ (*j,d*)/*N*_*surv*_ (*d*).

The distribution of symptomatic cases and hospitalisations by serotype, baseline serostatus and trial arm; by age group, baseline serostatus, and trial arm; and by serotype and age group are all assumed to follow multinomial distributions (see SI Section 2.5 for details).

#### Model variants and sensitivity analyses

To explore whether the data supported varying parameters by baseline serostatus, serotype, outcome, and age group, we considered multiple simpler nested models than those outlined above. Model variants were compared visually and quantitatively via the log-likelihood and the expected log-predictive density, estimated using the Watanabe-Akaike information Criterion, to find the most parsimonious, biologically-motivated model which could reproduce the trial data. Additionally, we tested fitting our final model to simulated data to confirm the identifiability of our model parameters (see **SI Section 2.6** for details). Specifically, we sampled each model parameter 20 times from distributions wider or centred away from the priors (**Supplementary Table 2**). We also generated simulated case data to reconstruct the posterior distribution of the parameters which was compared to the true (known) parameter value. We also conducted sensitivity analysis on the enhancement assumption by testing a model variant with no vaccine associated enhancement in seronegatives (*L*_*ck*_ =0 for all vaccinated individuals). **SI Section 2.7** provides full details on the sensitivity analyses run.

#### Inferential framework

The model was fitted to the baseline seropositivity data (*N*_*sp*_) and number of VCD symptomatic (*N*_*symp*_) and hospitalised (*N*_*hosp*_) cases observed across the trial, stratified by baseline serostatus, age and serotype within a Bayesian framework. Parameter inference was carried out using the Hamiltonian Monte Carlo algorithm and the No-U-Turn sampler via Stan, a probabilistic programming language implemented via the RStan package^22^ (version 2.26.21) in R^23^ (version 4.2.2). Four chains were run for 10,000 iterations each, and we discarded the first 5,000 iterations as burn-in. Convergence was assessed visually and using the 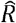 statistic^43^. The estimated parameters, priors and their sources are listed in **Supplementary Table 2**.

### Transmission Model

To evaluate the population-level impact of Qdenga, we applied the vaccine efficacy estimates obtained above to a 4-serotype stochastic compartmental model of dengue transmission, a deterministic version of which was published in Ferguson *et al*.^3^ (**Supplementary Figure 1b**). The model includes mosquito population dynamics with a seasonally varying carrying capacity. It models primary to quaternary human infections, assuming that homotypic immunity is lifelong and heterotypic immunity is temporary (12 months). It tracks the serotype-specific infection history but not the order of past infections. The model is stratified by vaccine status, serostatus, and age, with single age classes for the first 20 years and 10 years thereafter. See the **SI Section 3** for full details of the model. The model incorporates realistic non-stationary human demography, calibrated to the demographic estimates for Brazil and the Philippines published in the UN World Population Prospects 2022^44^. For this study, we embedded the VE model detailed above into this transmission model.

We explore four hypotheses about vaccine mechanism of action, obtained by combining (a) two hypotheses on the type of protection offered by the vaccine - only against disease (VS), parameterised from the survival model above, or also against infection (VI); (b) two assumptions for the period over which VE would wane, namely 5 years (approximately the period of the phase III trial) or 15 years (extrapolating beyond the trial). Full details on assumptions for the VI scenario are given **SI Section 2.3**. For each mechanism of action, we simulated the impact of routine vaccination of 6-to 12-year-olds across nine transmission intensity settings expressed in terms of average seroprevalence at nine years old (from 10% to 90% in steps of 10%). We assumed that all serotypes have near-identical transmissibility. We varied the vaccination coverage from 20% to 80% in steps of 20%. For each transmission setting, coverage scenario, and vaccine mechanism of action, we drew 200 samples from the posterior distribution of the parameters of the VE model and ran 50 stochastic simulations for each such posterior sample, giving 10,000 simulations per scenario in total (**Supplementary Figure 1b**). We summarise uncertainty in two ways. The first is the overall uncertainty, calculated over the 10,000 realisations, which represents uncertainty in both the posterior parameter estimates and the stochastic simulations. The second is the parameter uncertainty, calculated as the uncertainty of the mean of the 50 stochastic simulations for each of the 200 posterior samples.

To estimate the impact of vaccination, we compare the disease burden of each model run with a counterfactual simulation where we assume zero VE. Notably, for each realisation we matched exactly the counterfactual and vaccine scenario (i.e. the dengue dynamics up to the point of vaccine introduction are the same). We define the individual-level impact of vaccination as the proportion of cases averted in the first vaccinated cohort, and the population-level impact as the proportion of cases averted in the entire population (including non-vaccinated individuals) (**Supplementary Figure 1b**) after 10 years of vaccination, which we assumed started from 2024.

## References

1. Halstead, S. & O’Rourke, E. Dengue viruses and mononuclear phagocytes. I. Infection enhancement by non-neutralizing antibody. J. Exp. Med. 146, 201–217 (1977).

2. Sridhar, S. et al. Effect of dengue serostatus on dengue vaccine safety and efficacy. N. Engl. J. Med. 379, 327–340 (2018).

3. Ferguson, N. M. et al. Benefits and risks of the Sanofi-Pasteur dengue vaccine: Modeling optimal deployment. Science 353, 1033–1036 (2016).

4. Katzelnick, L. C. et al. Antibody-dependent enhancement of severe dengue disease in humans. Science 358, 929–932 (2017).

5. Salje, H. et al. Reconstruction of antibody dynamics and infection histories to evaluate dengue risk. Nature 557, 719–723 (2018).

6. Katzelnick, L. C., Montoya, M., Gresh, L., Balmaseda, A. & Harris, E. Neutralizing antibody titers against dengue virus correlate with protection from symptomatic infection in a longitudinal cohort. Proc. Natl. Acad. Sci. 113, 728–733 (2016).

7. Bos, S. et al. Protection against symptomatic dengue infection by neutralizing antibodies varies by infection history and infecting serotype. Nat. Commun. 15, 382 (2024).

8. Salje, H. et al. Variability in dengue titer estimates from plaque reduction neutralization tests poses a challenge to epidemiological studies and vaccine development. PLoS Negl. Trop. Dis. 8, 8–10 (2014).

9. Moodie, Z. et al. Neutralizing antibody correlates analysis of tetravalent dengue vaccine efficacy trials in Asia and Latin America. J. Infect. Dis. 217, 742–753 (2018).

10. Salje, H. et al. Evaluation of extended efficacy of Dengvaxia vaccine against symptomatic and subclinical dengue infection. Nat. Med. 27, 1395–1400 (2021).

11. Vannice, K. S. et al. Clinical development and regulatory points for consideration for second-generation live attenuated dengue vaccines. Vaccine 36, 3411–3417 (2018).

12. Alves, L. Brazil to start widespread dengue vaccinations. The Lancet 403, 133 (2024).

13. Osorio, J. E., Partidos, C. D., Wallace, D. & Stinchcomb, D. T. Development of a recombinant, chimeric tetravalent dengue vaccine candidate. Vaccine 33, 7112–7120 (2015).

14. Biswal, S. et al. Efficacy of a tetravalent dengue vaccine in healthy children and adolescents. N. Engl. J. Med. 381, 2009–2019 (2019).

15. Biswal, S. et al. Efficacy of a tetravalent dengue vaccine in healthy children aged 4–16 years: a randomised, placebo-controlled, phase 3 trial. The Lancet 395, 1423–1433 (2020).

16. López-Medina, E. et al. Efficacy of a dengue vaccine candidate (TAK-003) in healthy children and adolescents 2 years after vaccination. J. Infect. Dis. 225, 1521–1532 (2022).

17. Rivera, L. et al. Three-year efficacy and safety of Takeda’s dengue vaccine candidate (TAK-003). Clin. Infect. Dis. 75, 107–117 (2022).

18. Tricou, V. et al. Long-term efficacy and safety of a tetravalent dengue vaccine (TAK-003): 4·5-year results from a phase 3, randomised, double-blind, placebo-controlled trial. Lancet Glob. Health 12, e257–e270 (2024).

19. White, L. J. et al. Defining levels of dengue virus serotype-specific neutralizing antibodies induced by a live attenuated tetravalent dengue vaccine (TAK-003). PLoS Negl. Trop. Dis. 15, e0009258 (2021).

20. Waickman, A. T. et al. Assessing the diversity and stability of cellular immunity generated in response to the candidate live-attenuated dengue virus vaccine TAK-003. Front. Immunol. 10, (2019).

21. Tricou, V. et al. Characterization of the cell-mediated immune response to Takeda’s live-attenuated tetravalent dengue vaccine in adolescents participating in a phase 2 randomized controlled trial conducted in a dengue-endemic setting. Vaccine 40, 1143–1151 (2022).

22. WHO. WHO Position Paper on Dengue Vaccines – May 2024. 203–224 https://iris.who.int/bitstream/handle/10665/376641/WER9918-eng-fre.pdf (2024).

23. WHO. Meeting of the Strategic Advisory Group of Experts on Immunization, September 2023: Conclusions and Recommendations. https://iris.who.int/bitstream/handle/10665/374327/WER9847-eng-fre.pdf?sequence=1 (2023).

24. Khoury, D. S. et al. Neutralizing antibody levels are highly predictive of immune protection from symptomatic SARS-CoV-2 infection. Nat. Med. 2021 277 27, 1205–1211 (2021).

25. El Hindi, T. et al. Estimated efficacy of TAK-003 against asymptomatic dengue infection in children and adolescents. (2023).

26. Medina, F. A. et al. Comparison of the Sensitivity and Specificity of Commercial Anti-Dengue Virus IgG Tests to Identify Persons Eligible for Dengue Vaccination. 2024.04.19.24306097 Preprint at 10.1101/2024.04.19.24306097 (2024).

27. WHO SAGE. Background Paper on Dengue Vaccines. https://terrance.who.int/mediacentre/data/sage/SAGE_eYB_Sept2023.pdf (2023).

28. Osorio, J. E. et al. Safety and immunogenicity of a recombinant live attenuated tetravalent dengue vaccine (DENVax) in flavivirus-naive healthy adults in Colombia: a randomised, placebo-controlled, phase 1 study. Lancet Infect. Dis. 14, 830–838 (2014).

29. PAHO. PAHO Technical Advisory Group (TAG) on Immunization provides regional recommendations on vaccines against dengue, respiratory syncytial virus, and issues statement on ongoing use of COVID-19 vaccines - PAHO/WHO. https://www.paho.org/en/news/11-1-2024-paho-technical-advisory-group-tag-immunization-provides-regional-recommendations (2023).

30. Laydon, D. J. et al. Efficacy profile of the CYD-TDV dengue vaccine revealed by Bayesian survival analysis of individual-level phase III data. eLife 10, e65131 (2021).

31. Dorigatti, I. et al. Refined efficacy estimates of the Sanofi Pasteur dengue vaccine CYD-TDV using machine learning. Nat. Commun. 9, 3644 (2018).

32. Flanagan, K. L., Burl, S., Lohman-Payne, B. L. & Plebanski, M. The challenge of assessing infant vaccine responses in resource-poor settings. Expert Rev. Vaccines 9, 665–674 (2010).

33. Tappero, J. W. et al. Immunogenicity of 2 serogroup B outer-membrane protein meningococcal vaccines. A randomized controlled trial in Chile. JAMA 281, 1520–1527 (1999).

34. Cattarino, L., Rodriguez-Barraquer, I., Imai, N., Cummings, D. A. T. & Ferguson, N. M. Mapping global variation in dengue transmission intensity. Sci. Transl. Med. 12, 1–11 (2020).

35. WHO. Dengue vaccine: WHO position paper, September 2018 – Recommendations. Vaccine 37, 4848–4849 (2019).

36. Rodríguez-Barraquer, I., Salje, H. & Cummings, D. A. Dengue pre-vaccination screening and positive predictive values. Lancet Infect. Dis. 19, 132–134 (2019).

37. Thommes, E. et al. Public health impact and cost-effectiveness of implementing a ‘pre-vaccination screening’ strategy with the dengue vaccine in Puerto Rico. Vaccine 40, 7343–7351 (2022).

38. St. John, A. L. & Rathore, A. P. S. Adaptive immune responses to primary and secondary dengue virus infections. Nat. Rev. Immunol. 19, 218–230 (2019).

39. Kallás, E. G. et al. Live, attenuated, tetravalent Butantan–dengue vaccine in children and adults. N. Engl. J. Med. 390, 397–408 (2024).

40. Utarini, A. et al. Efficacy of wolbachia-infected mosquito deployments for the control of dengue. N. Engl. J. Med. 384, 2177–2186 (2021).

41. Goethals, O. et al. Blocking NS3–NS4B interaction inhibits dengue virus in non-human primates. Nature 615, 678–686 (2023).

42. Carpp, L. N. et al. Microneutralization assay titer correlates analysis in two phase 3 trials of the CYD-TDV tetravalent dengue vaccine in Asia and Latin America. PLOS ONE 15, e0234236 (2020).

43. Gelman, A. & Rubin, D. B. Inference from Iterative Simulation Using Multiple Sequences. Stat. Sci. 7, 457–472 (1992).

44. United Nations. World Population Prospects. (2022).

