## Supplementary Information for "Efficacy, public health impact and optimal use of the Takeda dengue vaccine"

### Supplementary methods

#### 1. Data

Per protocol (subjects who received both doses with no major protocol violations), outcomes and population sizes for the Qdenga phase III clinical trial are shown in **Supplementary Figure 2.** The number of symptomatic virologically confirmed dengue (VCD) cases, $N_{symp}$, were published by trial arm, $v$ (= 1 for placebo, = 2 for vaccinated), classified baseline serostatus $b$ (- = seronegative, + = seropositive), and infecting serotype $k$ (=1, 2, 3, 4 for DENV1-4 respectively), within each reporting interval $d$ (1= 1-12 months, 2 = 13-18 months, 3 = 19-24 months, 4 = 25-36 months, 5 = 37-48 month, 6 = 49-54 months). The number of hospitalised cases, $N_{hosp}$, were published by trial arm, baseline serostatus and infecting serotype at 1-24 months (safety set, individuals who received at least one dose of the vaccine or placebo), as well as for the last three-reporting intervals. $N_{symp}$ and $N_{hosp}$ were both published by trial arm, baseline serostatus and age group, $j$ (1 = 4-5yrs, 2= 6-11yrs, 3 = 12-16yrs), for the first four reporting intervals $d$ (*i.e.*, up to 36 months), and by age group and infecting serotype for reporting intervals 1-12 months and 13-24 months. Limited country-specific data was published, so all data and model estimates presented here are for the whole trial population across all countries.

The trial populations by baseline serostatus and trial arm were reported for each time interval, $N_{pop_{bv}}(d)$, with the population further broken down by age group $j$ up to 36 months (**Supplementary Figure 2G and 2H**). Population sizes decreased over the duration of the trial, as participants were lost to follow-up. As only the first VCD case is reported in the trial, we right-censor cases at the end of each reporting interval $d$ and reduce the monitored population in the next time interval accordingly (*i.e.*, $N_{surv_{bv}}(j,d) = N_{pop_{bv}}(j,d) - N_{symp_{bv}}(j,d-1)$ ). Age-specific data were not published after month 36 of the trial, so we estimate the age-specific trial populations at 37-48 months and 49-54 months, assuming the same age-specific probability of loss to follow-up as the first 36 months of the trial.

#### 2. Cohort survival model

**2.1 Probability of exposure at baseline:**
We estimate the probability of exposure to each serotype $k$ in age group 3 (12-16 years) prior to the start of the trial $(t = T_0)$, $p_{k3}$. For the other two age groups, we estimate the cumulative probabilities of exposure to any serotype and scale serotype-specific probabilities to match the distribution of serotype-specific exposure in age group 3. Let $h_{k3}$ be the serotype-specific hazard of exposure in age group 3:

$$h_{k3} = -\ln(1 - p_{k3}) \tag{1}$$

The serotype-specific hazards in age groups $j = 1$ and $j = 2$ are therefore given by:

$$h_{kj} = \frac{h_{k3}}{\sum_k h_{k3}} * -\ln\left(1 - p_j\right) \tag{2}$$

Finally, the serotype-specific probabilities of exposure in age groups $j = 1$ and $j = 2$ are:

$$p_{kj} = 1 - e^{-h_{kj}} \tag{3}$$

To aid identifiability of both the FOI and probability of symptoms, we constrain the probability of past exposure $p$ by the estimated FOI during the trial. Let $\lambda_m$ be the average monthly FOI during the trial (irrespective of serotype). The cumulative probability of exposure in the oldest age group $p_m$ is therefore:

$$p_m = 1 - e^{(-14*12*\lambda_m)} \tag{4}$$

where 14 is the mean age in the oldest age group and the factor of 12 reflects the time unit of months used in our analysis. We therefore set a beta prior for $p_{k3}$ with a mean $p_m$ and variance $var = 0.02 * p_m(1 - p_m)$. A value of 0.02 was chosen to ensure the prior was informative. We reparametrised the beta distribution to get standard shape parameters $shape1$ and $shape2$.

##### 2.2 Misclassification of serostatus at baseline:

Accounting for the imperfect test sensitivity, $sens$, and specificity, $spec$, of the microneutralisation test used to classify individuals as seronegative or seropositive at baseline, the probability of being seronegative at baseline $pE_{\emptyset}(j, T_0)$ (defined in the Methods section of the main text) and classified as seronegative ($b = -$) is:

$$pE_{\emptyset -}(j, T_0) = spec * pE_{\emptyset}(j, T_0) \tag{5}$$

The probability of being seronegative at baseline and misclassified as seropositive ($b = +$) is:

$$pE_{\emptyset +}(j, T_0) = (1 - spec) * pE_{\emptyset}(j, T_0) \tag{6}$$

The probability of having had prior exposure to serotypes in set Ω at baseline, $pE_{\Omega}(j, T_0)$, and being misclassified as seronegative is:

$$pE_{\Omega -}(j, T_0) = (1 - sens) * pE_{\Omega}(j, T_0) \tag{7}$$

The probability of having had prior exposure at baseline and being classified as seropositive is:

$$pE_{\Omega +}(j, T_0) = sens * pE_{\Omega}(j, T_0) \tag{8}$$

##### 2.3 Vaccine efficacy for hospitalised dengue

The antibody titre-based model of vaccine efficacy (VE) used in our analysis is presented in the Methods section of the main text. Supplementing the description provided there, the risk of being hospitalised with dengue for a participant in the vaccination arm of the trial relative to one in the control arm is:

$$\mathrm{RR}_{hosp_{cvkj}}(t_i) = \frac{1 + \tau_k * L_{ck}}{1 + \left(\frac{n_{ck}(t_i)}{e^{-\alpha_{ck}} * e^{\beta_{hosp_j}} * n_{50_{ck3}}}\right)^{w_{ck}}} \quad if\ v = 2 \tag{9}$$

This is the same model as used for the symptomatic VCD endpoint (see equation 4 in Methods section of main text) with two modifications: (a) the maximum level of vaccine-induced enhancement for symptomatic VCD, $L_{ck}$, is scaled by the multiplier $\tau_k$; (b) the neutralising titre which gives 50% protection from hospitalisation, $n_{50}$ is assumed to be proportional to that for VCD, but scaled by the factor $e^{-\alpha_{ck}}$. In addition, the variation of efficacy with age (represented by the parameter $\beta_{hosp_j}$) is allowed to be different from that for the symptomatic VCD endpoint. Other parameters ($w_{ck}$, $n_{50_{ck3}}$ and $L_{ck}$) are as defined as in equation 4 in the main text.

##### 2.4 Incidence of infection, symptomatic disease, hospitalisation

The incidence of primary infection is:

$$Inc_{\Omega bvk}(j, t) = \left(1 - \mathrm{e}^{-\lambda_k(t)}\right) pE_{\emptyset bv}(j, t) \tag{10}$$

where $\lambda_k(t)$ is the serotype-specific FOI in month $t$ and $pE_{\emptyset bv}(j, t)$ is the probability of being seronegative.

The incidence of secondary infection is:

$$Inc_{\Omega bvk}(j, t) = \left(1 - \mathrm{e}^{-\lambda_k(t)}\right) \sum_k pE_{\Omega_{\setminus k} bv}(j, t) \tag{11}$$

where $pE_{\Omega_{\setminus k} bv}(j, t)$ is the probability of having prior exposure to serotypes in set $\Omega \in h_1 = \{\{1\}, \{2\}, \{3\}, \{4\}\}$, not including serotype $k$.

The incidence of tertiary infection is:

$$Inc_{\Omega bvk}(j, t) = \left(1 - \mathrm{e}^{-\lambda_k(t)}\right) \sum_k pE_{\Omega_{\setminus k} bv}(j, t) \tag{12}$$

where $\Omega \in h_2 = \{\{1,2\},\{1,3\},\{1,4\},\{2,3\},\{2,4\},\{3,4\}\}$.

The incidence of quaternary infection is:

$$Inc_{\Omega bvk}(j,t) = \left(1 - \mathrm{e}^{-\lambda_k(t)}\right) \sum_k pE_{\Omega \backslash \mathrm{k} bv}(j,t) \tag{13}$$

where $\Omega \in h_3 = \{\{1,2,3\},\{1,2,4\},\{1,3,4\},\{2,3,4\}\}$

The incidences of primary, secondary, and post-secondary disease are respectively given by:

$$Symp_{\Omega bvk}(j,t) = \rho_k \gamma \, \mathrm{RR}_{symp_{0vkj}}(t) \, Inc_{\Omega bvk}(j,t) \tag{14}$$

$$Symp_{\Omega bvk}(j,t) = \gamma \, \mathrm{RR}_{symp_{1vkj}}(t) \, Inc_{\Omega bvk}(j,t) \tag{15}$$

$$Symp_{\Omega bvk}(j,t) = \rho_k \varphi \gamma \, \mathrm{RR}_{symp_{2vkj}}(t) \left(Inc_{\Omega bvk}(j,t)\right) \tag{16}$$

where $\Omega \in h_1$ in equation 14, $\Omega \in h_2$ in equation 15 and $\Omega \in \{h_3, h_4\}$ in equation 16 and $\gamma$ is the probability of symptoms during a secondary infection, $\rho_k$ is the risk of symptoms during a primary infection relative to a secondary infection, $\varphi$ is the risk of symptomatic disease during a post-secondary infection compared to a primary infection and $\mathrm{RR}_{symp_{1vkj}}$ is the vaccine associated risk ratio of symptomatic disease as defined in equation 4 in the Methods section of the main text.

The incidences of primary, secondary, and post-secondary hospitalisation are:

$$Hosp_{\Omega bvk}(j,t) = \delta_k \, \rho_k \, \gamma \, \mathrm{RR}_{hosp_{0vk}}(t) \, Inc_{\Omega bvk}(j,t) \tag{17}$$

$$Hosp_{\Omega bvk}(j,t) = \delta_k \epsilon \, \gamma \, \mathrm{RR}_{hosp_{1vk}}(t) \, Inc_{\Omega bvk}(j,t) \tag{18}$$

$$Hosp_{\Omega bvk}(j,t) = \delta_k \rho_k \varphi \gamma \, \mathrm{RR}_{hosp_{2vk}}(t) \left(Inc_{\Omega bvk}(j,t)\right) \tag{19}$$

where $\Omega \in h_1$ in equation 17, $\Omega \in h_2$ in equation 18 and $\Omega \in \{h_3, h_4\}$ in equation 19 and $\delta_k$ is the probability that a symptomatic case due to serotype $k$ is hospitalised and $\epsilon$ is the risk of hospitalisation in secondary cases, compared primary or post-secondary infections.

The total symptomatic and hospitalisation incidences are:

$$Symp_{bvk}(j,t) = \sum_{\Omega \in \{h_1, \mathrm{h}_2, \mathrm{h}_3, \mathrm{h}_4\}} Symp_{\Omega bvk}(j,t) \tag{20}$$

$$Hosp_{bvk}(j,t) = \sum_{\Omega \in \{h_1, \mathrm{h}_2, \mathrm{h}_3, \mathrm{h}_4\}} Hosp_{\Omega bvk}(j,t) \tag{21}$$

**2.5 Multinomial Likelihood**

For each published time interval d, the expected distribution of symptomatic cases ($pS_{bvk}(d)$) or hospitalisations ($pH_{bvk}(d)$) in baseline serostatus $b$, due to serotype $k$, in trial arm $v$, relative to the total number of symptomatic cases ($cumS(d) = \sum_b \sum_v \sum_k \sum_j S_{bvk}(j,d)$) or hospitalisations ($cumH(d) = \sum_b \sum_v \sum_k \sum_j H_{bvk}(j,d)$), are given by:

$$pS_{bvk}(t_d) = \frac{\sum_j S_{bvk}(j,d)}{cumS(d)} \tag{22}$$

$$pH_{bvk}(t_d) = \frac{\sum_j H_{bvk}(j,d)}{cumH(d)} \quad (23)$$

Equally, $pS_{bv}(j,d)$ and $pH_{bv}(j,d)$, the expected proportion of symptomatic cases and hospitalisations in baseline serostatus $b$, trial arm $v$, age group $j$ are given by:

$$pS_{bv}(j,d) = \frac{\sum_k S_{bvk}(j,d)}{cumS(d)} \quad (24)$$

$$pH_{bv}(j,d) = \frac{\sum_k Hosp_{bvk}(j,d)}{cumH(d)} \quad (25)$$

Finally, $pS_k(j,d)$ and $pH_k(j,d)$, the expected proportion of symptomatic cases or hospitalisations due to serotype $k$ in age group $j$ are given by:

$$pS_k(j,d) = \frac{\sum_b \sum_v S_{bvk}(j,d)}{cumS(d)} \quad (26)$$

$$pH_k(j,d) = \frac{\sum_b \sum_v Hosp_{bvk}(j,d)}{cumH(d)} \quad (27)$$

The expected probabilities of observing symptomatic disease and hospitalised cases given in eqs. 25-30 are assumed to follow multinomial distributions:

$$N_{symp_{bvk}}(d) \sim Multinomial\left(pS_{bvk}(d)\right) \quad (28)$$

$$N_{symp_{bv}}(j,d) \sim Multinomial\left(pS_{bv}(j,d)\right) \quad (29)$$

$$N_{symp_k}(j,d) \sim Multinomial\left(pS_k(j,d)\right) \quad (30)$$

$$N_{Hosp_{bvk}}(d) \sim Multinomial\left(pH_{bvk}(d)\right) \quad (31)$$

$$N_{Hosp_{bv}}(j,d) \sim Multinomial\left(pH_{bv}(j,d)\right) \quad (32)$$

$$N_{Hosp_k}(j,d) \sim Multinomial\left(pH_k(j,d)\right) \quad (33)$$

##### 2.6 Simulation study to assess the model identifiability

To confirm the identifiability of the model parameters given the data, we conducted a simulation study. We sampled 20 sets of the parameters used to generate simulated datasets from distributions that were wider or centred away from the prior distributions used for parameter inference. The only exception was the parameters describing the titre decay ($hs$, $hl$, and $ts$), which were estimated directly from the observed data and were intentionally kept informative. We then simulated case data using each set of parameters. The case data were comparable to the observed data in magnitude and distribution across time, age group, serostatus, and trial arm. Finally, we calibrated the survival model to the simulated case data and compared the simulated posterior distribution of each parameter to the true parameter distributions.

##### 2.7 Model sensitivity analyses

We ran a sensitivity analysis on the choice of the $L_{ck}$ prior normal distribution centred and truncated at 0, with standard deviations {0.25, 0.5, 0.75, 1.00, 1.25, 1.50, 1.75, 2.00}. We assessed the impact of the choice of the prior distribution on the log-likelihood, posterior estimate of $L_{ck}$, and the Bayes factor estimate comparing models with and without vaccine associated enhancement (i.e., setting $L_{ck}$ = 0).

We also explored the sensitivity of the VE estimates to assumptions made about (i) the period of heterotypic cross-immunity, which we fixed at 1 and 6 months (rather than 12 months as in our main analysis), (ii) whether multitypic individuals could be misclassified as seronegative at baseline, and (iii) whether all post-secondary infections are asymptomatic.

#### 3. Transmission model

##### 3.1 Mosquito dynamics

The transmission model accounts for mosquito population dynamics following the Ross-MacDonald model, with varying seasonal carrying capacity. Specifically, adult female mosquitoes lay eggs at rate $\Gamma$ and larvae ($L$) mature into adult mosquitos ($M$) at rate $\mathrm{E}$. Larvae are regulated by a power density dependent mortality rate $\mu$, with a seasonal carrying capacity $K$, ensuring that mortality increases as the larval population increases. Adult mosquitoes are born susceptible ($M_S$), experience a serotype-specific FOI $\Lambda_k$, and infected mosquitoes enter an exposed but not infectious compartment ($M_E$) which lasts on average for a period $\frac{1}{\xi}$ days, the extrinsic incubation period. Finally, the adult mosquitoes that survive the extrinsic incubation period become infectious to humans ($M_I$) until their death. Adult mosquitoes die at a rate $\Delta$. The ordinary differential equations (ODE) governing the mosquito dynamics are given as follows:

$$\frac{dL}{dt} = \Gamma M - (\mathrm{E} + \mu) L$$
$$\frac{dM_S}{dt} = \mathrm{E}\, L - \left(\Delta + \sum_k \Lambda_k\right) M_S$$
$$\frac{dM_{E_k}}{dt} = \Lambda_k\, M_S - (\xi + \Delta) M_{E_k}$$
$$\frac{dM_{I_k}}{dt} = \xi\, M_{E_k} - \Delta\, \mathrm{M}_{\mathrm{I}_k} \tag{34}$$

The mortality rate $\mu$ of the larvae is defined as:

$$\mu = \sigma\left(1 + \frac{L}{K\, N}\right) \tag{35}$$

Where $\sigma$ is the low-density limit of larval death and $N$ is the total human population.

In turn, the seasonal carrying capacity $K$ is defined as:

$$K = \overline{K}(1 + K_s \cos(2\pi t * 365)) \tag{36}$$

Where $\overline{K}$ is the mean carrying capacity, $K_s$ is the magnitude of the seasonal variation in carrying capacity, and $t$ is time in days. The mean carrying capacity is calculated to match the observed adult wild-type female mosquito density per person, $\mathrm{A}$:

$$\overline{K} = \mathrm{A}\, \Delta\, \frac{\left(\mathrm{E}\, \frac{\Gamma - \Delta}{\Delta\sigma} - 1\right)^{-\frac{1}{\xi}}}{\mathrm{E}} \tag{37}$$

The rate at which adult female mosquitoes lay eggs $\Gamma$ is assigned to give the required mosquito population reproduction number (i.e., representing the reproductive capacity of the mosquito population, rather than anything to do with disease transmission):

$$R_{0_m} = \Delta\, \frac{\mathrm{E}}{\mathrm{E} + \mu}\, \Gamma \tag{38}$$

The probability of transmission from human to mosquito $\mathrm{B}_{hm}$ is defined to give the required dengue $R_0$ as follows:

$$\mathrm{B}_{hm} = \frac{R_0}{\kappa^2 \, \Lambda \, \eta \frac{\frac{\mathrm{B}_{mh}}{1+\Delta\,\xi}}{\Delta}} \tag{39}$$

Where $\eta$ is the human infectious period and $\kappa$ is the biting rate per mosquito.

Finally, the force of infection experienced by mosquitoes due to dengue serotype $k$, $\Lambda_k$ is:

$$\Lambda_k = \mathrm{B}_{hm} \, \kappa \frac{cI_k}{N} \tag{40}$$

Where $cI_k$ is the human incidence of infection (defined below in equation 47) with serotype $k$ and $N$ is the total size of the human population.

##### 3.2 Human transmission dynamics

As with the cohort survival model, let subscript $\Omega$ denote the set of serotypes an individual has previously been exposed to (with cardinality $c$). In the transmission model, we assume that individuals are born susceptible to all four serotypes $S_{\emptyset}$, after which they are subject to serotype-specific FOI $\lambda_k(t)$. Infection with any of the four serotypes confers long-lasting homotypic immunity and heterotypic immunity against the other serotypes for a period $1/\vartheta$ (12 months) ($R_{\Omega}$). After this, individuals are assumed to be susceptible to infection with heterotypic serotypes. In total, an individual may be infected up to four times, and we track an individual's infection history but not the serotype-specific order of infection.

The model is additionally stratified by the age group $i$ (1 to 20 years of single age classes, then 10-year age classes up to 80 years), and vaccination status $v$ (1 = unvaccinated, 2 = vaccinated), with individuals aged $f$ vaccinated with coverage $\upsilon$. Aging is modelled as an annual discrete event (e.g. all 1 year-olds moving to the 2 year-old age group) to track the vaccinated cohort accurately up to the age of 20. We assume that vaccination also occurs once per year at the same time as the ageing process, and we do not separately model receiving the first and second dose. The birth rate $\Psi(\mathrm{t})$ and the age-specific human mortality rate $\theta_{\mathrm{i}}(t)$ are both calibrated to match the demography of either the Philippines or Brazil (see section **3.4** below for additional details). The model is solved daily ($t$). The ODEs governing human transmission dynamics are:

$$\frac{dS_{\Omega vi}}{dt} = \delta_{\Omega,\emptyset}\delta_{\mathrm{v},1}\delta_{\mathrm{i},0}\Psi(t)S_{\Omega vi} + \delta_{\mathrm{i},f}\,\upsilon\left[\delta_{\mathrm{v},2}\,S_{\Omega vi} - \delta_{\mathrm{v},1}\,S_{\Omega vi}\right] - \left(\sum_{k\notin\Omega}\lambda_{\mathrm{k}}(t)\,\mathrm{RR}_{{inf}_{cvki}}(t) + \theta_{\mathrm{i}}(t)\right)S_{\Omega vi}$$

$$\frac{dR_{\Omega vi}}{dt} = \delta_{\mathrm{i},f}\,\upsilon\left[\delta_{\mathrm{v},2}\,R_{\Omega vi} - \delta_{\mathrm{v},1}\,R_{\Omega vi}\right] + \left(\sum_{k\notin\Omega}\lambda_{\mathrm{k}}(t)\mathrm{RR}_{{inf}_{cvki}}(t)\right)S_{\Omega vi} - (\vartheta + \theta_{\mathrm{i}}(t))R_{\Omega vi} \tag{41}$$

Here $\delta_{\mathrm{x,y}}$ is the Kronecker delta function (equal to 1 when $\mathrm{x} = \mathrm{y}$ and 0 otherwise), the first term of $\frac{dS_{\Omega vi}}{dt}$ represents births, the second term represents vaccination (which occurs once a year at the same time as ageing) and the final term represents infections and deaths. The parameter $\mathrm{RR}_{{inf}_{cvki}}(t)$ is the vaccine-associated RR of infection (see section **3.4** below for details). The first term of $\frac{dR_{\Omega vi}}{dt}$ represents vaccination, the second term represent infection, and the third term represents waning heterotypic immunity and mortality. All transitions between compartments (ageing, infection, waning immunity, death, and vaccination) are drawn from binomial distributions to introduce model stochasticity).

The FOI on humans due to serotype $k$ is:

$$\lambda_{\mathrm{k}}(t) = \mathrm{B}_{mh}\kappa \frac{M_{I_k}}{N} \tag{42}$$

where $\mathrm{B}_{mh}$ is the per bite transmission probability from mosquitoes to humans.

The incidence of individuals exposed (but not yet infectious) to serotype $k$ is given as:

$$E_{cvki}(t) = \lambda_{k \notin \Omega}(t)\, \mathrm{RR}_{inf_{cvki}}(t)\, S_{\Omega vi} \tag{43}$$

The incidence of individuals who are infectious $I$, symptomatic $D$, or hospitalised $H$ due to serotype $k$ at time $t$ are given as:

$$I_{cvki}(t) = \zeta E_{cvki} \tag{44}$$

$$D_{cvki}(t) = \mathrm{RR}_{symp_{cvki}}(t)\, p_{symp_c}\, I_{cvki}(t) \tag{45}$$

$$H_{cvki}(t) = \mathrm{RR}_{hosp_{cvki}}(t)\, Q\, p_{symp_c}\, I_{cvki}(t) \tag{46}$$

Here $1/\zeta$ is the human incubation period, $p_{symp_c}$ is the probability an infected individual with serostatus $c$ is symptomatic (= $\rho\,\gamma$ for primary infections, $\gamma$ for secondary infections, and $\rho\,\gamma\,\varphi$ for post-secondary infections, see equations 14-16 above for parameter descriptions), $Q$ is the probability a symptomatic individual requires hospitalisation (fixed at 9% to match the average probability of hospitalisation observed in Brazil and the Philippines during the phase III clinical trial), and $\mathrm{RR}_{symp_{cvki}}(t)$ and $\mathrm{RR}_{hosp_{cvki}}(t)$ are the vaccine-associated RR of disease and hospitalisation (defined in equation 9 above and equation 4 of the main text).

The cumulative incidence of individuals infectious to serotype $k$, irrespective of serostatus, vaccine status, or age group, as seen in equation 40 above is therefore:

$$cI_k = \sum_c \sum_v \sum_i I_{cvki} \tag{47}$$

##### 3.3 Model parameter values

The VE parameters ($hs$, $hl$, $ts$, $L$, $n_{50}$, $\alpha$, $\beta$, $w$, and $\tau$) and probabilities of symptoms ($\rho$, $\gamma$, $\varphi$) were sampled from the posterior distribution of the survival model used to reconstruct $RR_{symp}$ and $RR_{hosp}$ and described in Section 2. All other parameter values used in the stochastic compartmental model of transmission are given in **Supplementary Table 3.**

##### 3.4 Model simulations

We equilibrated the transmission dynamics by running the transmission model in the absence of vaccination for 175 years, starting from 1850. For the first 100 years we assumed a statistic 1950's demography. From 1950 onwards we assumed time-varying demographies matching the UN World Population Prospects 2022 estimates for Brazil or the Philippines[1].

We ran multiple scenarios to investigate the vaccines impact across a) two demographies (Brazil and the Philippines); b) nine transmission intensity settings, defined as the average seroprevalence at 9-years-old, from 10% to 90% in steps of 10%; c) four vaccine coverages (20%, 40%, 60%, 80%); d) seven ages at vaccination (from 6 to 12 years of age); e) four hypotheses about the vaccines mechanism of action, obtained by assuming vaccine waning for either 5 years or 15 years post-vaccination, and that vaccine protects against clinical disease only (VS), or also against infection (VI); and f) with and without pre-vaccination screening of individuals serostatus assuming a diagnostic test with 94.7% specificity and 89.6% sensitivity[2].

In the absence of serological tests able to identify the infecting serotype and infer asymptomatic infections, data on Qdenga's efficacy against infection is limited, but some evidence suggests that it may offer some protection against infection for some months[3]. In the VI modelling scenarios we assumed that the probability of infection, as for clinical outcome, can be explained by the antibody titre induced by previous infection[2] or vaccination [3]. We assume that the titres required for protection from infection are higher than those required to protect against symptomatic disease, and that (unlike enhancement of disease) there is no enhancement of infection risk[4,5]. We therefore modelled a scenario of moderate VE against infection by scaling the posterior estimates for $n_{50_{ck3}}$(the neutralising titre conferring 50% protection against disease in the absence of infection) in eq. 4 of the main text by $\alpha_{\mathrm{inf}_c}$ , equal to 12 if $c = 0$ (primary infection) and 3 if $1 \le c \le 3$:

$$\mathrm{RR}_{inf_{cvkj}}(t) = \frac{1}{1 + \left(\frac{n_{ck}(t)}{\alpha_{\mathrm{inf}_c}\, e^{\beta_{symp_j}}\, n_{50_{ck3}}}\right)^{w_{ck}}} \qquad if\ v = 2 \qquad (48)$$

The resulting $\mathrm{VE}_{inf}$ curves are plotted in **Supplementary Figure 15.**

For each of the modelling scenarios, we sampled the VE parameters from the posterior distributions 200 times (**Supplementary Table 1**). Then, for each posterior parameter set we ran the stochastic transmission model 50 times (where the probability of infection and vaccination are drawn from binomial distributions), giving 10,000 simulations in total for each scenario (**Supplementary Figure 1b**).

Each model simulation was run twice: once with VE set to our estimated values ("vaccination" scenario) and once with VE set to zero ("no vaccination" scenario). This enabled us to estimate the population- and individual-level impact of vaccination. Individual impact is measured as the proportion of cases averted in the first vaccinated cohort (over 10 years) and population-level impact is the proportion of cases averted in the entire population (over 10 years) (**Supplementary Figure 1b**):

$$prop_{av} = \frac{(cases_{NV} - cases_V)}{cases_{NV}}$$

Here $cases_{NV}$ are the number of cases in the zero VE ("no vaccination") counterfactual scenario, and $cases_V$ are the number of cases in the VE>0 ("vaccination") scenario.

We estimated the overall uncertainty of the impact estimates as the 95% CrI across all 10,000 simulations, so that the interval accounts for the parameter uncertainty of the efficacy estimates and dynamic uncertainty of the stochastic simulations. We estimated the parameter uncertainty as the 95% CrI of the mean across the 50 stochastic simulations calculated across the 200 posterior estimates of the efficacy parameters.

We note that the vaccination and counterfactual/no vaccination runs were matched exactly (i.e., identical transmission dynamics were simulated for each such pair of simulations) up to the point of vaccine introduction. In the absence of any VE against infection, transmission dynamics remained identical for vaccination and no-vaccination scenarios following the introduction, but this was not the case if non-zero VE against infection was assumed (since vaccination then modified transmission).

### Supplementary results

#### Efficacy projections

**Supplementary Figure 10** shows the VE projections up to year 15 post-vaccination, assuming the same rate of efficacy decay estimated for the trial period. VE against both symptomatic and hospitalised dengue remains moderate to high against DENV2, especially for seronegative individuals. For the other serotypes, the vaccine is estimated to offer little or no protection against disease or hospitalisation in seropositive individuals by year 15. For seronegative individuals, the model predicts that by year 15, the vaccine would substantially enhance disease and hospitalisation following a DENV1, DENV3, and DENV4 infection, irrespective of age.

#### Sensitivity analyses

We compared our main model to one with no disease enhancement (fixing $L_{ck}$ = 0), given different standard deviations of the prior distribution (truncated normal, centred at 0). Although the magnitude of the Bayes factor estimate was sensitive to the choice of the prior distribution (range: 7-35), it consistently supported the model with enhancement (**Supplementary Figures 11a and 11b**). Notably, a standard deviation <1 restricted the posterior estimate of the enhancement parameter and worsened the model fit (**Supplementary Figures 11c and 11d**). We, therefore, chose a standard deviation of 1 for the main analysis. This gave us a Bayes factor of 13, implying strong but not irrefutable evidence of enhancement (**Supplementary Figure 11a**).

We ran sensitivity analyses to assess the impact of assuming (i) shorter periods of heterotypic cross-immunity, (ii) perfect classification of multitypic individuals as seropositive at baseline and (iii) no symptomatic post-secondary infections. The VE estimates obtained in these sensitivity analyses were similar to those obtained in the main model (**Supplementary Figure 12**).

### Supplementary tables

**Supplementary Table 1:** Mean and 95% CrI posterior parameter estimates of the survival model (M30), used to estimate vaccine efficacy.

| Parameter | | | Estimate (mean and 95% CrI) |
|---|---|---|---|
| $hs_c$ | Seronegative | | 1.93 (1.12 to 2.72) |
| | Seropositive | | 4.34 (3.58 to 5.11) |
| $hl$ | - | | 72.14 (49.38 to 94.25) |
| $ts_c$ | Seronegative | | -2.10 (-2.93 to -1.28) |
| | Seropositive | | 0.31 (-0.52 to 1.13) |
| $p_j$ | 4-5yrs | | 0.23 (0.21 to 0.24) |
| | 6-11yrs | | 0.30 (0.28 to 0.31) |
| | 12-16yrs | | 0.47 (0.44 to 0.50) |
| $\gamma$ | - | | 0.46 (0.34 to 0.63) |
| $1/\rho$ | - | | 2.44 (2.13 to 2.79) |
| $\varphi$ | - | | 0.26 (0.18 to 0.34) |
| $\delta_k$ | DENV1 | | 0.19 (0.15 to 0.23) |
| | DENV2 | | 0.40 (0.34 to 0.47) |
| | DENV3 | | 0.17 (0.12 to 0.21) |
| | DENV4 | | 0.14 (0.06 to 0.24) |
| $L$ | - | | 0.84 (0.13 to 1.84) |
| $\tau_k$ | DENV1 | | 1.31 (0.22 to 2.57) |
| | DENV2 | | 1.26 (0.14 to 2.70) |
| | DENV3 | | 2.18 (0.81 to 3.55) |
| | DENV4 | | 1.14 (0.12 to 2.55) |
| $w$ | - | | 2.73 (1.80 to 3.89) |
| $lc_{50_{ck}}$ | Seronegative | | 4.86 (4.46 to 5.28) |
| | Monotypic | DENV1 | 6.74 (6.54 to 6.92) |
| | Monotypic | DENV2 | 6.98 (6.58 to 7.28) |
| | Monotypic | DENV3 | 6.82 (6.61 to 7.04) |
| | Monotypic | DENV4 | 6.04 (5.48 to 6.58) |
| | Multitypic | | 6.51 (4.91 to 7.81) |
| $\alpha$ | - | | 0.49 (0.30 to 0.74) |
| $\beta_j$ | 4-5yrs | | 0.49 (0.29 to 0.76) |
| $sens$ | - | | 0.91 (0.89 to 0.93) |
| $spec$ | - | | 0.99 (0.98 to 1.00) |
| $\lambda_k\ (t_d)$ | DENV1 to DENV4 | 1-12, 13-18, 19-24, 25-36, 37-48, and 49-54 months | Range: $6.32^{-05}$ ($5.22^{-06}$ to $1.93^{-04}$) - $1.15^{-02}$ ($7.16^{-03}$ to $1.69^{-02}$) |

**Supplementary Table 2: Survival model parameters and their priors.** To aid convergence, $n_{50_{ck3}}$ is estimated on the log scale, $lc_{50_{ck3}}$.

| Parameter | Description | Prior distribution | Source |
|---|---|---|---|
| $\boldsymbol{hs_c}$ | Short half-life of antibody decay in seronegatives (c= $0$). | Normal (1.65,0.5) | [9] |
| $\boldsymbol{hs_c}$ | Short half-life of antibody decay in seropositives ($c = 1 = 2 = 3$). | Normal (4.20,0.50) | [9] |
| $\boldsymbol{hl}$ | Long half-life of antibody decay. | Normal (84.00,12.00) | [25] |
| $\boldsymbol{ts_c}$ | Time period switch decay in seronegatives ($c = 0$). | Normal (-2.21, 0.50) | [9] |
| $\boldsymbol{ts_c}$ | Time period switch decay in seropositives ($c = 1 = 2 = 3$). | Normal (0.15, 0.50) | [9] |
| $\boldsymbol{L_{ck}}$ | Enhancement of symptomatic disease in vaccinated seronegative individuals ($c = 0$) | Normal (0.00,1.00) | |
| $\boldsymbol{L_{ck}}$ | Enhancement of symptomatic disease in vaccinated seropositive individuals ($c = 1 = 2 = 3$). | Fixed at 0.00 | |
| $\boldsymbol{w_{ck}}$ | Shape parameter. | Normal (1.00,2.00) | [26] |
| $\boldsymbol{lc_{50_{ckj}}}$ | Log neutralising antibody titre against each serotype which provides 50% protection against symptomatic disease for seronegative individuals in age group 3 ($c = 0, j = 3$). | Normal (4.50,1.00) | [9,16-18] |
| $\boldsymbol{lc_{50_{ckj}}}$ | Log neutralising antibody titre against each serotype which provides 50% protection against symptomatic disease for seropositive individuals in age group 3 ($c = 1 = 2 = 3, j = 3$). | Normal (6.50,1.00) | [9,16-18] |
| $\boldsymbol{\tau_k}$ | Risk ratio of enhancement of hospitalisation compared to enhancement of symptomatic disease. | Normal (1.00,1.00) | |
| $\boldsymbol{\alpha_{ck}}$ | Difference in neutralising antibody titre required for 50% protection from hospitalisation compared to symptomatic disease. | Normal (0.00,2.00) | |
| $\boldsymbol{\beta_{symp_j}}$ | Difference in neutralising antibody titre required for 50% protection from symptomatic disease in age groups 1 and 2 ($j = 1, 2$), compared to age group 3. | Normal (0.00,2.00) | |
| $\boldsymbol{\beta_{hosp_j}}$ | Difference in neutralising antibody titre required for 50% protection from hospitalisation in age groups 1 and 2 ($j = 1,2$), compared to age group 3. | Normal (0.00,2.00) | |
| $\boldsymbol{p_j}$ | Probability of exposure in age groups 1 and 2 ($j = 1, 2$). | Beta (3.00,5.00) | |
| $\boldsymbol{p_{kj}}$ | Probability of exposure to serotype $k$ in age group 3 ($j = 3$). | Beta ($shape1, shape2$) | |
| $\boldsymbol{\lambda_k(t_d)}$ | Serotype-specific force of infection during the trial. | Lognormal (-7.00,2.00) | |
| $\boldsymbol{\gamma}$ | Probability a secondary infection is symptomatic. | Normal (0.85,0.20) | [27] |
| $\boldsymbol{1/\rho_k}$ | Risk ratio of disease in primary infections compared to secondary infections. | Normal (1.97,0.40) | [27] |
| $\boldsymbol{\varphi}$ | Probability that a post-secondary infection is symptomatic compared to a primary infection. | Normal (0.25,0.05) | [27] |
| $\boldsymbol{\delta_k}$ | Probability a primary or post-secondary symptomatic case is hospitalised. | Normal (0.25,0.10) | [28,29] |
| $\boldsymbol{\epsilon}$ | Risk ratio of hospitalisation in secondary cases compared to primary or post-secondary cases. | Normal (1.00,1.00) | |

| | | | |
|---|---|---|---|
| $\boldsymbol{sens}$ | Sensitivity of the microneutralisation test used to classify serostatus at baseline. | Normal (0.90,0.05) | [19] |
| $\boldsymbol{spec}$ | Specificity of the microneutralisation test used to classify serostatus at baseline. | Normal (0.995,0.01) | [19] |

**Supplementary Table 3: Transmission model parameters.**

| Parameter | Description | Value | Source |
|---|---|---|---|
| $\boldsymbol{\Gamma}$ | Rate at which adult females produce female larvae | Calculated to give the required $R_{0_m}$ | |
| $\boldsymbol{\bar{K}}$ | Mean larval mosquito carrying capacity | Calculated to match the required adult wild-type female mosquito density per person. | |
| $\boldsymbol{\mu}$ | Larval mosquito mortality rate | Derived from the larvae dependence in eq. 41 | |
| $\boldsymbol{\xi}$ | Extrinsic incubation period | 8 days | |
| $\boldsymbol{\Delta}$ | Adult mosquito death rate | 0.1 | [6] |
| $\mathbf{E}$ | Rate larvae develop into adult mosquitoes | Assigned 1/19 days, to match $R_{0_m}$ | [6] |
| $\boldsymbol{\sigma}$ | Low-density limit of larval death | 0.025 | [6] |
| $\boldsymbol{K_s}$ | Magnitude of the seasonal variation in carrying capacity | 0.3 | [6] |
| $\mathbf{A}$ | Adult wild-type female mosquito density per person | 1.5 | [6] |
| $\boldsymbol{R_{0_m}}$ | | 2.69 | [6] |
| $\mathbf{B_{mh}}$ | Probability of transmission from mosquito to human | 1.0 | [6] |
| $\mathbf{B}_{\boldsymbol{hm}}$ | Probability of transmission from human to mosquito | Calculated to give the required reproduction number in eq. 38 | |
| $\boldsymbol{R_0}$ | Reproduction number | Varied from ~1.1 to ~5 to match required equilibrium pre-vaccination seropositivity in 9-year-olds of 10-90%. | |
| $\boldsymbol{\eta}$ | Human infectious period | 4 days | [6] |
| $\boldsymbol{\kappa}$ | Biting rate per mosquito per day | 0.6 | [6] |
| $\boldsymbol{\vartheta}$ | Period of heterotypic immunity | 12 months | [6] |
| $\boldsymbol{\theta_i}$**(t)** | Human mortality rate | Fitted to either Brazilian or Philippines demography from the UN World Population Prospects | [1] |
| $\boldsymbol{\Psi(t)}$ | Human birth rate | Scaled estimates from the UN World Population Prospects corresponding to Brazilian or Philippines to give a population of 5 million in 1950 | |
| $\boldsymbol{\zeta}$ | Intrinsic incubation period | 5 days | |
| $\boldsymbol{\upsilon}$ | Vaccination coverage | Varied: 20%, 40%, 60%, 80% | |
| $\boldsymbol{Q}$ | Proportion of symptomatic infections which require hospitalisation in Brazil or the Philippines | Set as 0.09 to match the average probability of hospitalisation observed in Brazil and the Philippines during the phase III clinical trial | |

#### Supplementary figures

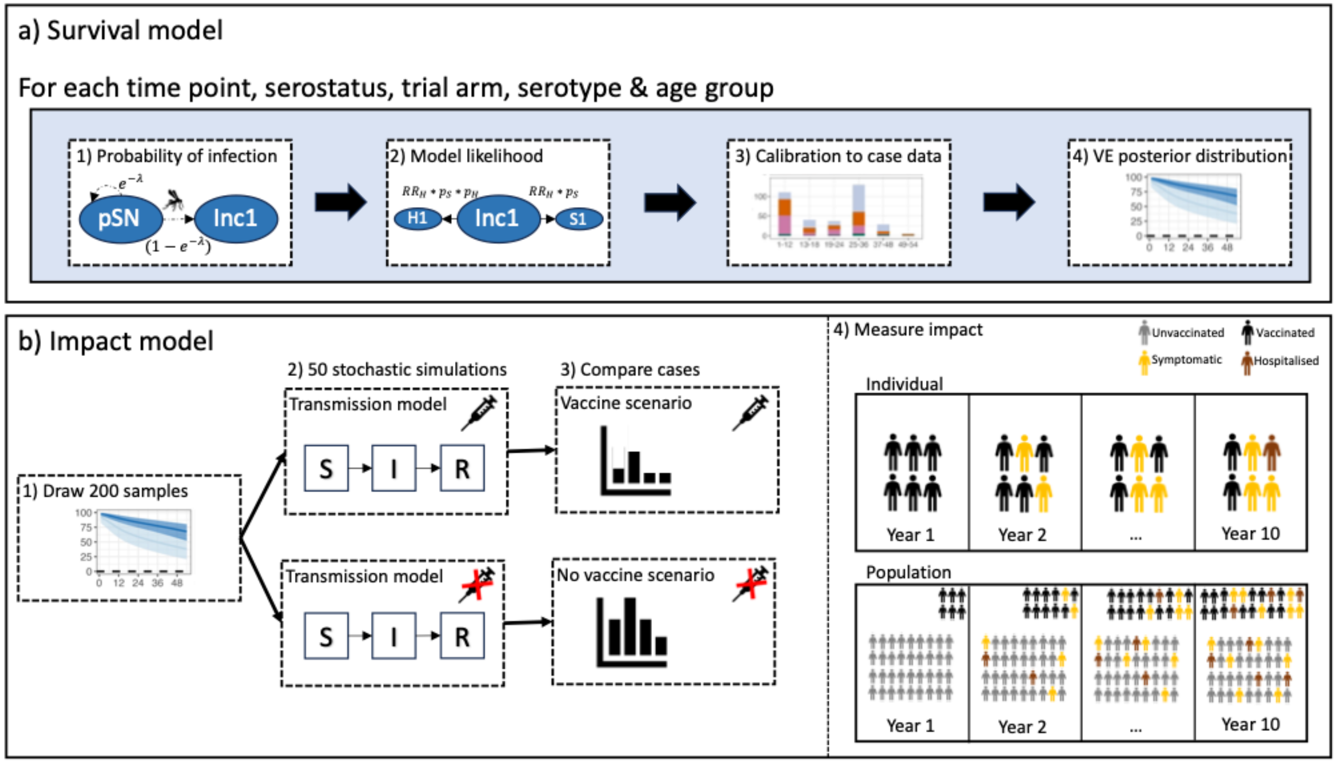

**Supplementary Figure 1: Conceptual figure of the modelling approach using in the study. (a)** Overview of the Bayesian cohort survival model used to reproduce the phase III clinical trial case data and estimate vaccine efficacy, using a seronegative individual as an example and outlining how the model keeps track of (1) the probability that seronegative individuals (pSN) are infected and (2) the incidences of primary symptomatic disease (S1) and hospitalisation (H1) are reconstructed from the incidence of primary infection (Inc1) through the vaccine-associated risk ratios of symptomatic disease and hospitalisation $RR_S$ and $RR_H$ , respectively and (3) the calibration of the survival model to the published case data, which allows (4) the estimation of the vaccine efficacy (VE) over-time. **(b)** Overview of the modelling framework used to estimate the impact of routine vaccination with the compartmental model. For each scenario of routine vaccination, we drew 200 posterior samples of the VE parameters and for each sample, we (2) run 50 simulations of the compartment transmission model with (top) and without (bottom) vaccination, to output (3) the expected number of cases and thus (4) assess the impact of vaccination in two ways: at the individual-level (top) impact is calculated as the proportion of cases averted in the first vaccinated cohort over ten years; at the population-level (bottom) impact is calculated as the proportion of cases averted in the entire population (including the non-vaccinated population) over ten years of routine vaccination.

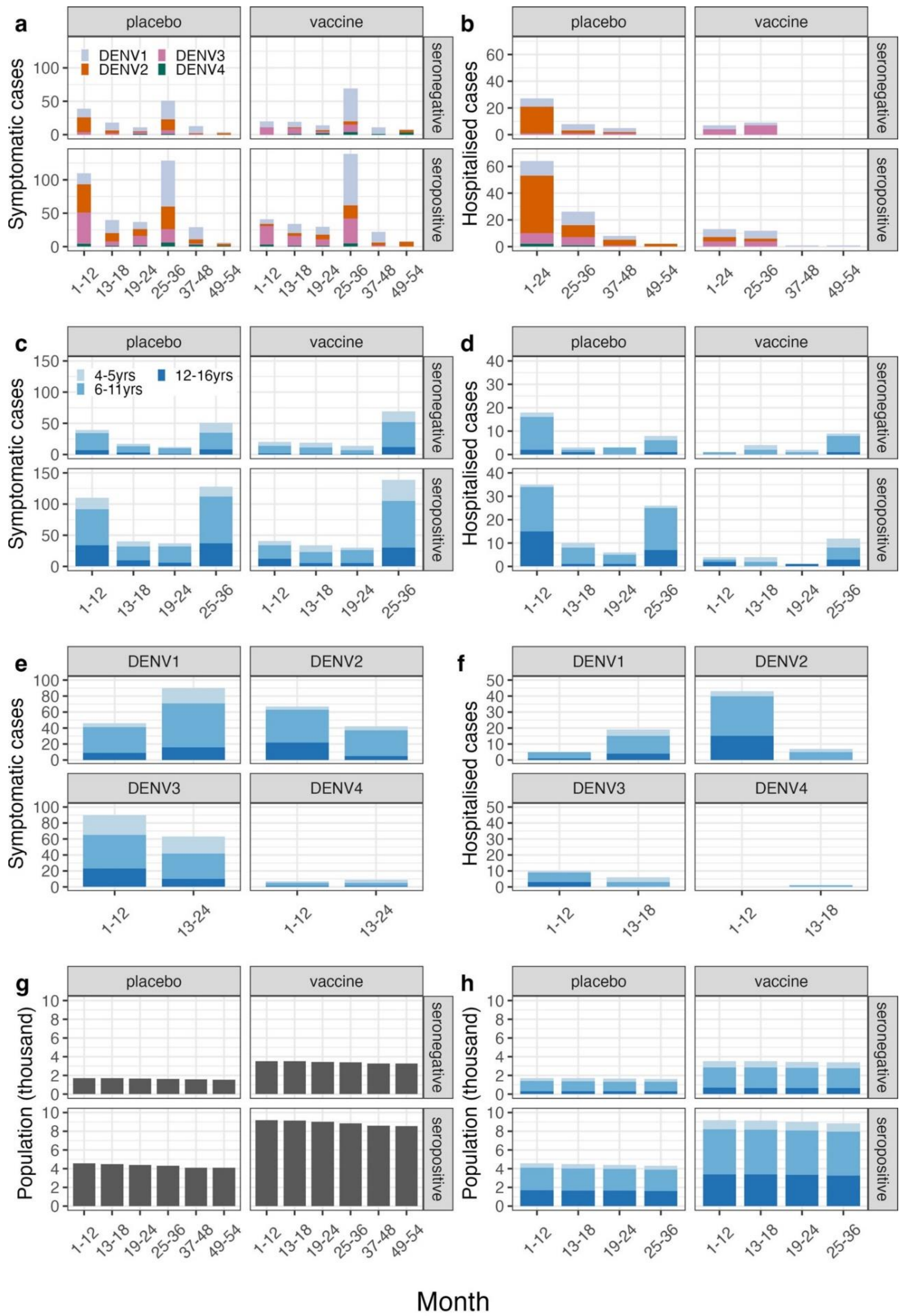

**Supplementary Figure 2: Summary of the trial data used to calibrate the model. (a, c, e) Symptomatic cases. (b, d, f) Hospitalised cases. (g-h) Population sizes.** Note the 2:1 randomisation of individuals to receive the vaccine vs. the placebo. Data are from the per-protocol population (individuals without any major protocol violations, including not receiving both doses of the correct assignment of Qdenga or placebo) except for the serotype-specific hospitalisation case data in months 1-24 (**b**) which is from the safety population (individuals who received at least one dose of the vaccine or placebo).

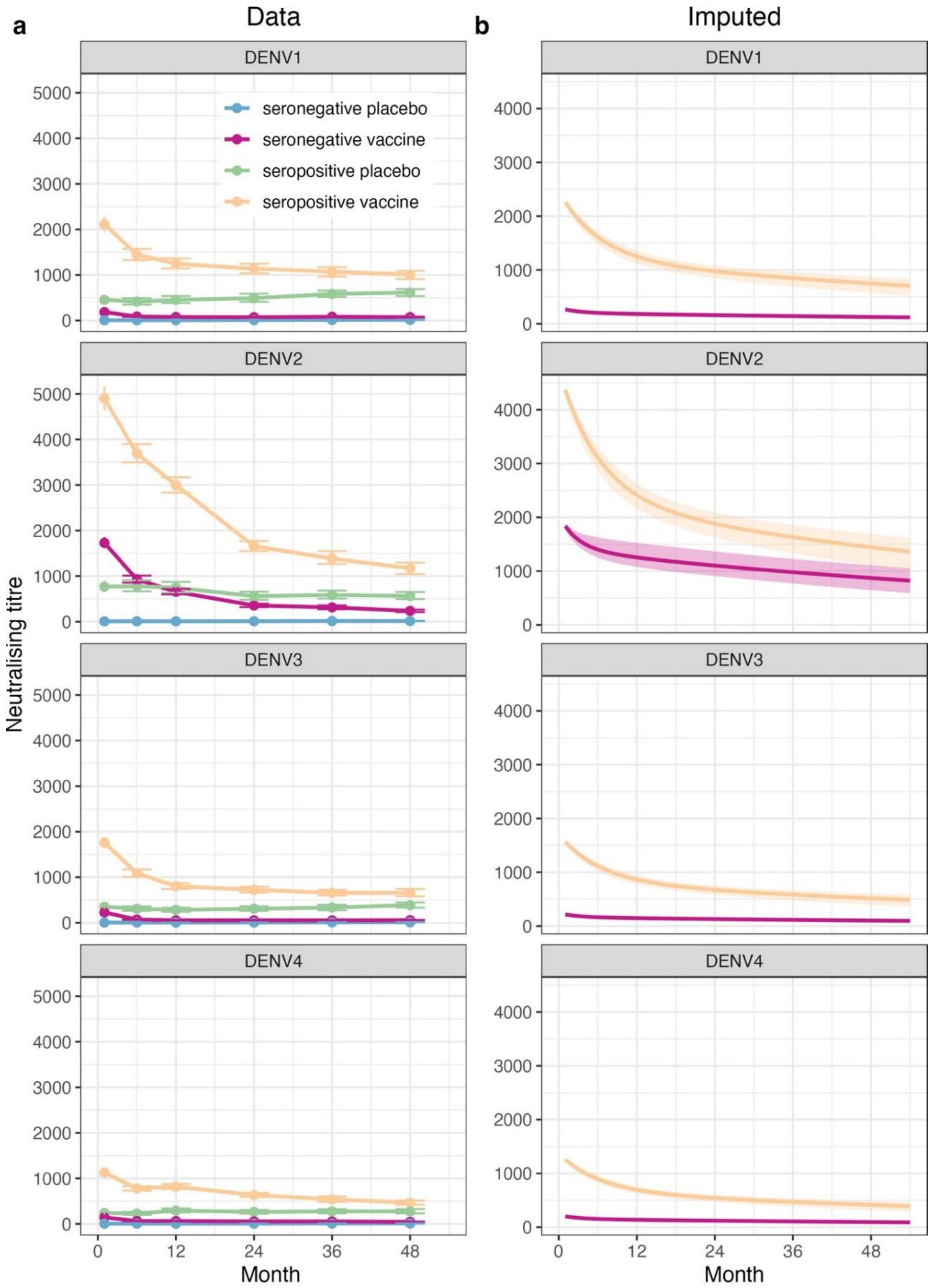

**Supplementary Figure 3: Neutralising antibody titres. (a)** Observed mean (point) and 95% confidence interval (error bar) neutralising antibody titres by serostatus and trial arm (colours) against each serotype (rows). **(b)** Imputed mean (solid line) and 95% credible interval (shaded region) of the neutralising antibody titres in the vaccine trial arm by serostatus (colours) and serotype (rows).

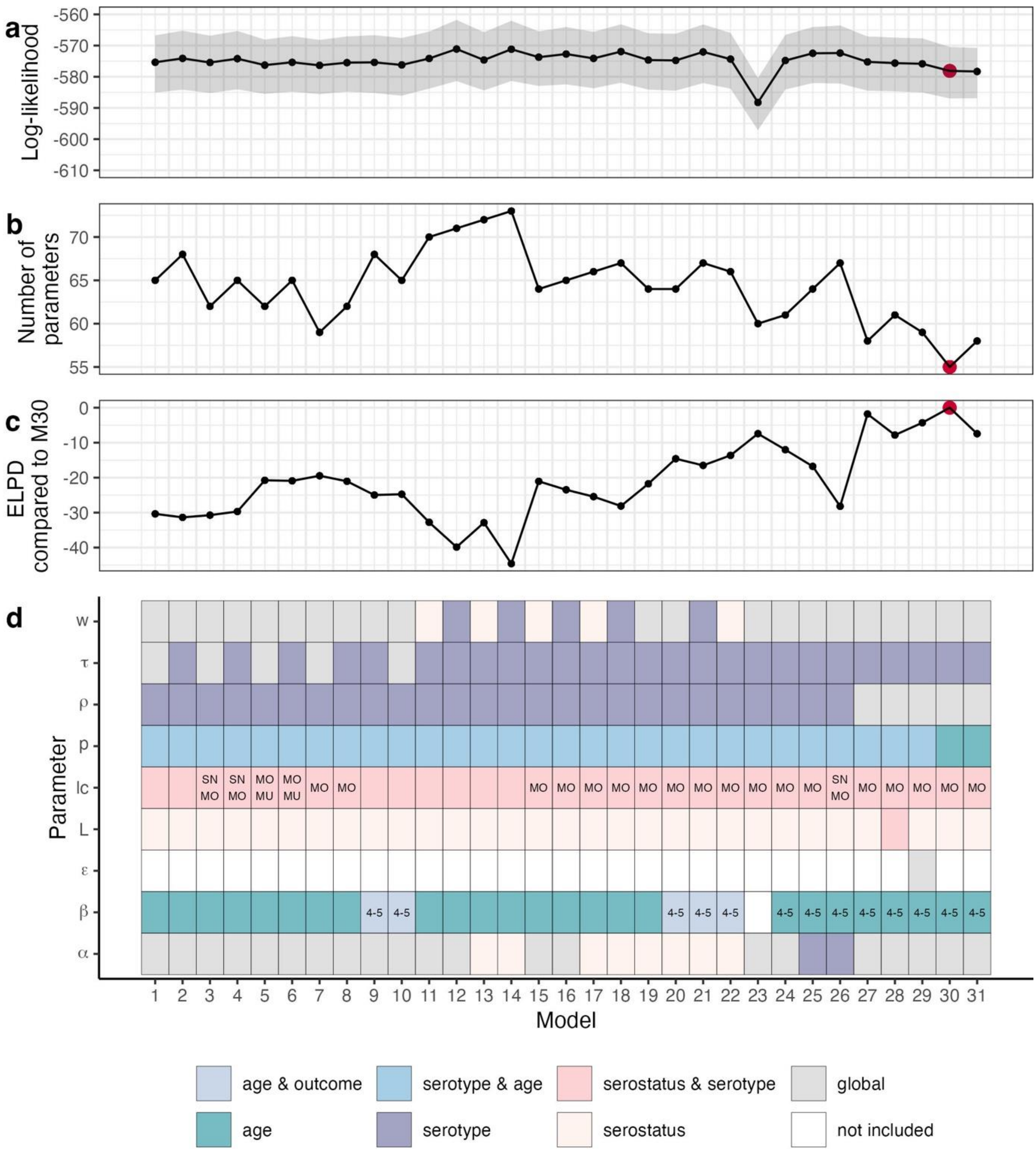

**Supplementary Figure 4: (a) Log-likelihood, (b) number of parameters, (c) expected log predictive density (ELPD) compared to the main model and (d) visual description of the estimated parameters by model variant.** See Supplementary Table 2 for a full description of the model parameters. The final model (30) is shown in red.

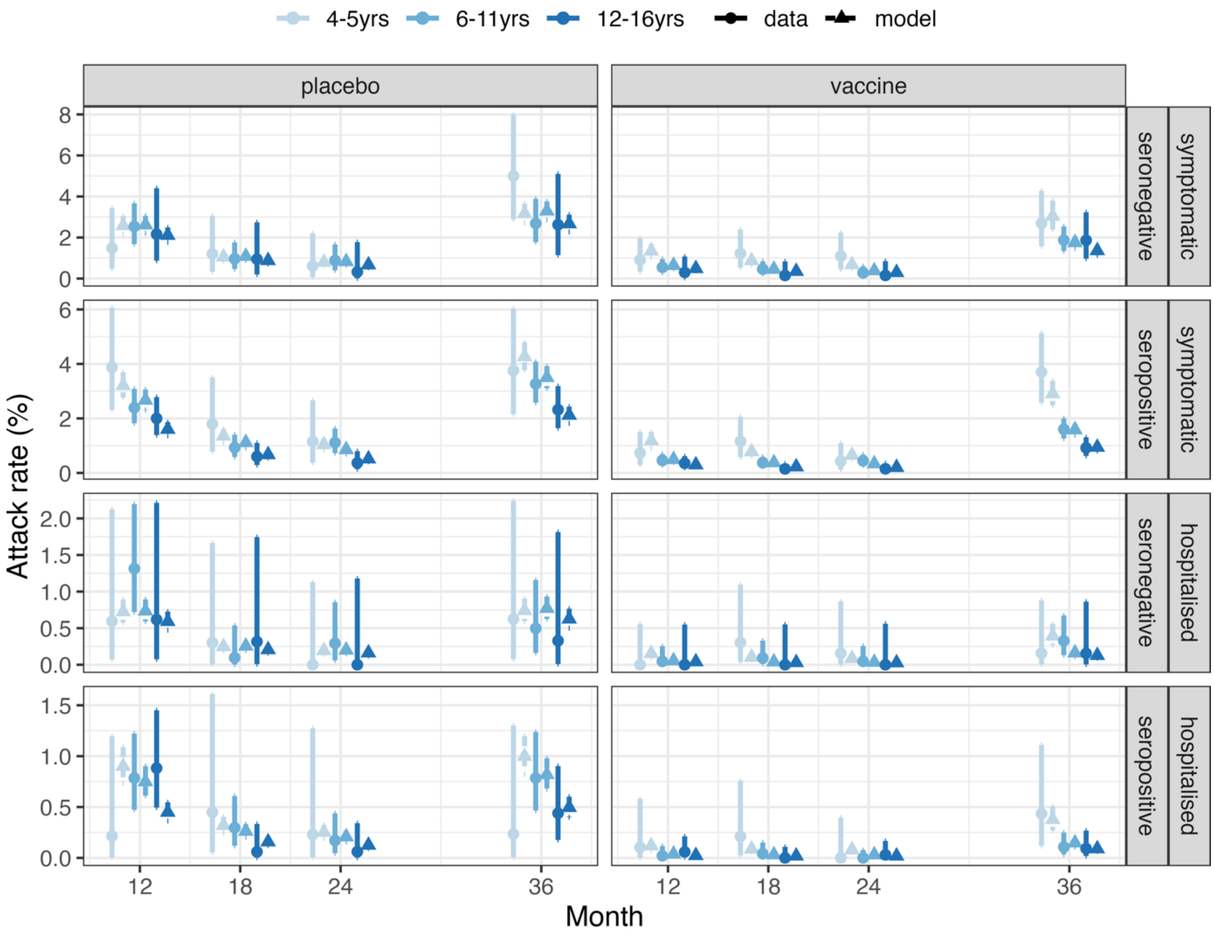

**Supplementary Figure 5: Model calibration to age-specific case data.** Observed and estimated symptomatic and hospitalised attack rates during the phase III clinical trial by trial arm, serostatus, age group, and time. The modelled attack rates show the mean (triangle) and 95% credible interval (dashed line). The observed attack rates show the mean (circle) and 95% exact binomial confidence interval (solid line).

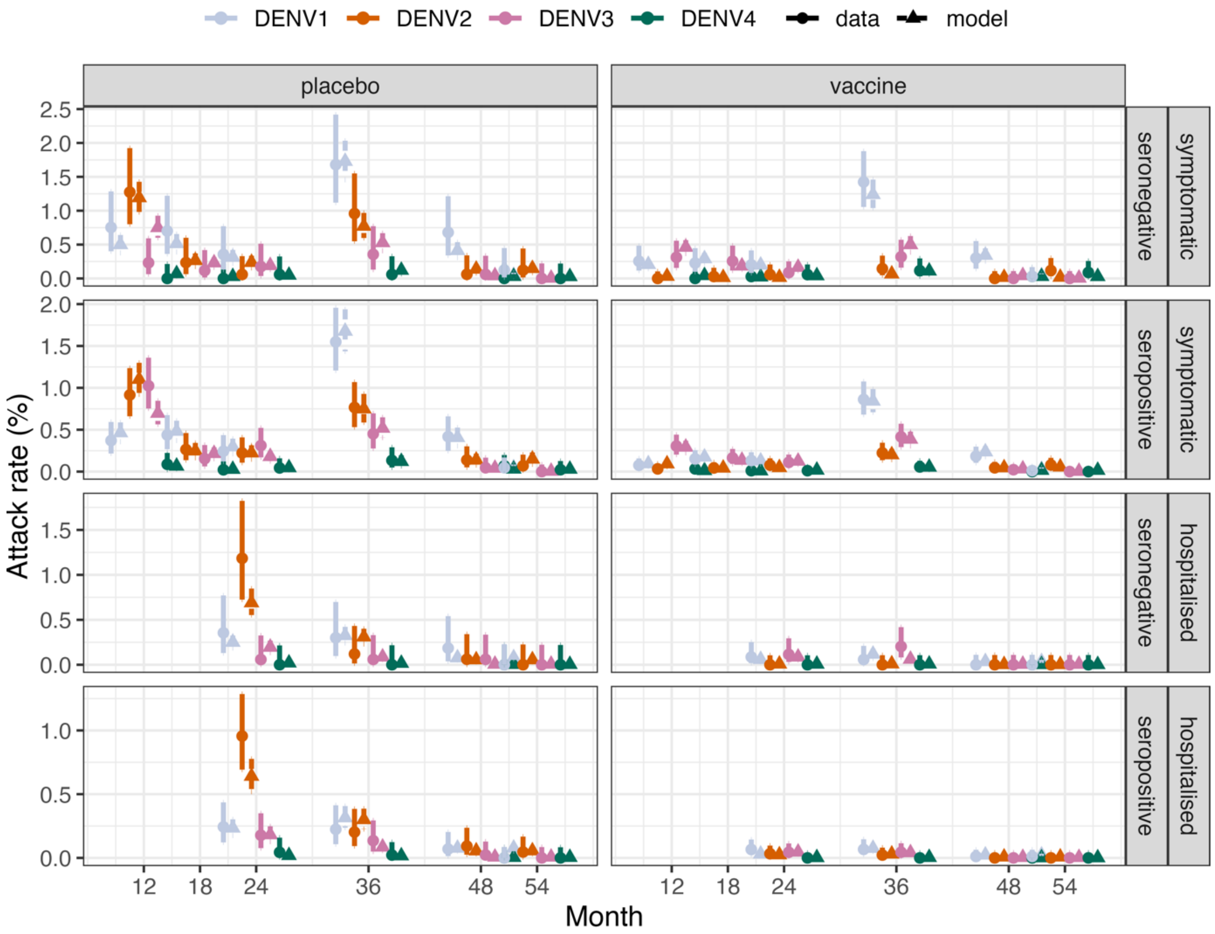

**Supplementary Figure 6: Model calibration to serotype-specific case data.** Observed and estimated symptomatic and hospitalised attack rates during the phase III clinical trial by serostatus, trial arm, serotype, and time. The modelled attack rates show the mean (triangle) and 95% credible interval (dashed line). The observed attack rates show the mean (circle) and 95% exact binomial confidence interval (solid line).

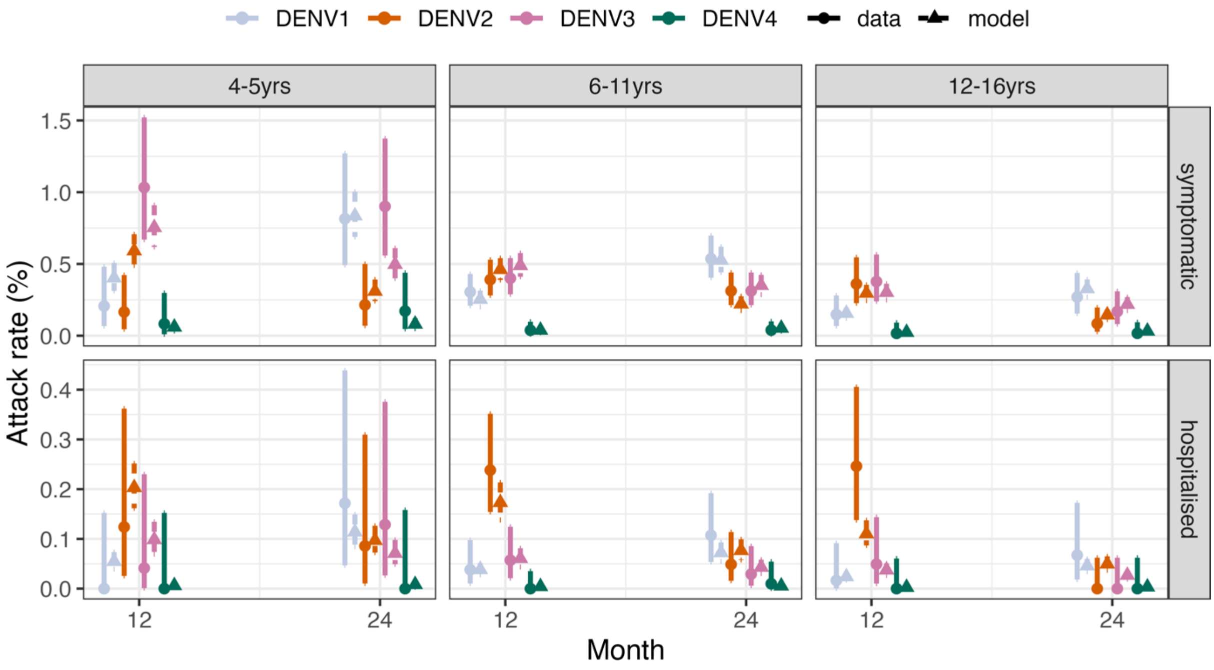

**Supplementary Figure 7: Model calibration to age- and serotype-specific case data.** Observed and estimated symptomatic and hospitalised attack rates during the phase III clinical trial by serotype, age group, and time. The modelled attack rates show the mean (triangle) and 95% credible interval (dashed line). The observed attack rates show the mean (circle) and 95% exact binomial confidence interval (solid line).

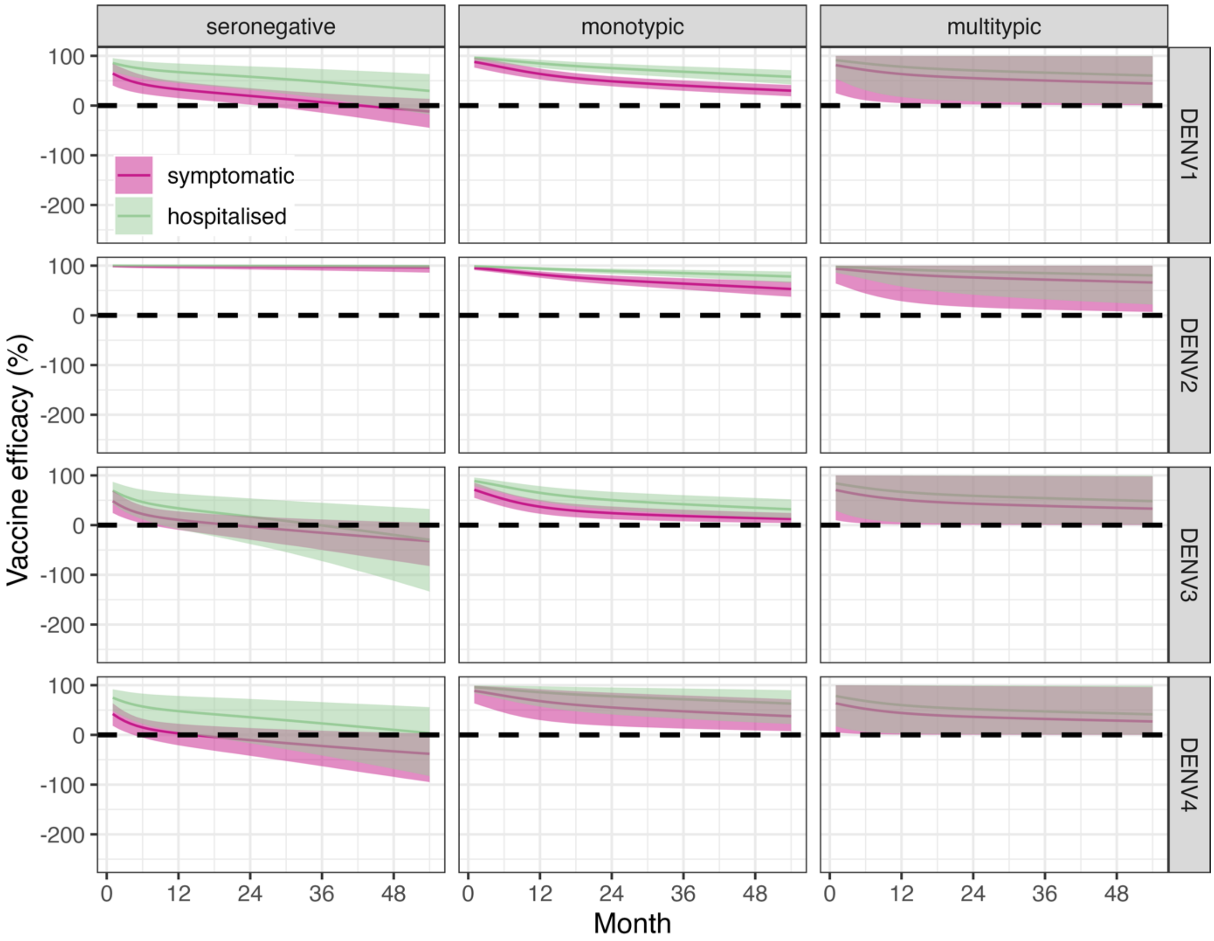

**Supplementary Figure 8: Vaccine efficacy estimates.** Estimated vaccine efficacy by serostatus (columns) and serotype (rows), against symptomatic disease and hospitalisation (colours). The solid line represents the mean efficacy and the shaded area represents the 95% credible interval. The dashed horizontal line marks 0 efficacy.

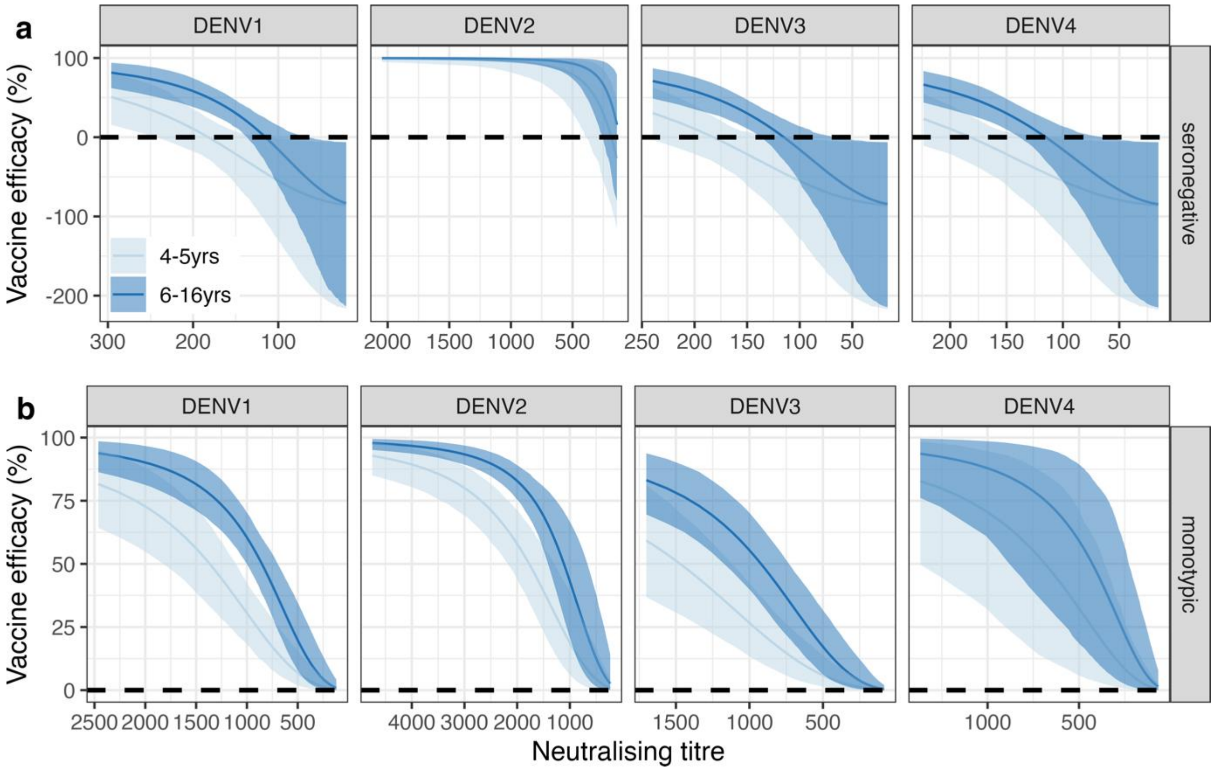

**Supplementary Figure 9: Relationship between vaccine efficacy and neutralising antibody titre.** Vaccine efficacy by serotype (columns), age (colours) against symptomatic disease in **(a)** seronegative individuals and **(b)** monotypic individuals. The solid line represents the mean efficacy and the shaded area represents the 95% credible interval. The dashed horizontal line marks 0 efficacy. Neutralising titres are projected to 20 years post-vaccination assuming the same decay rate observed during the phase III clinical trial. Note the different x-axis scales.

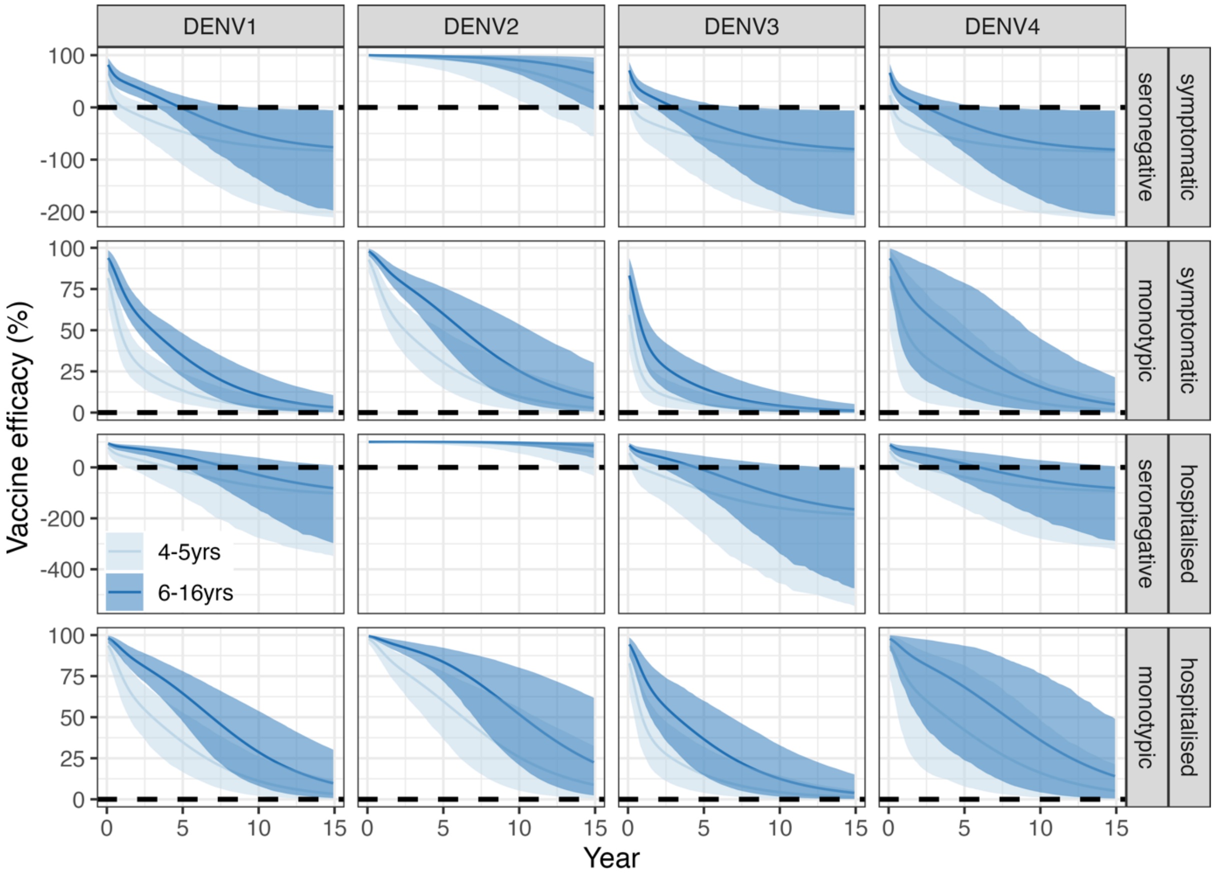

**Supplementary Figure 10: Vaccine efficacy estimates projected up to 15 years post-vaccination.** Vaccine efficacy by serotype (columns), serostatus (rows) and age (colours) against symptomatic disease and hospitalisation (rows). The solid line represents the mean efficacy and the shaded area represents the 95% credible interval. The dashed horizontal line marks 0 efficacy. Efficacy estimates assume the same decay rate observed during the phase III clinical trial. Note the different y-axis scales.

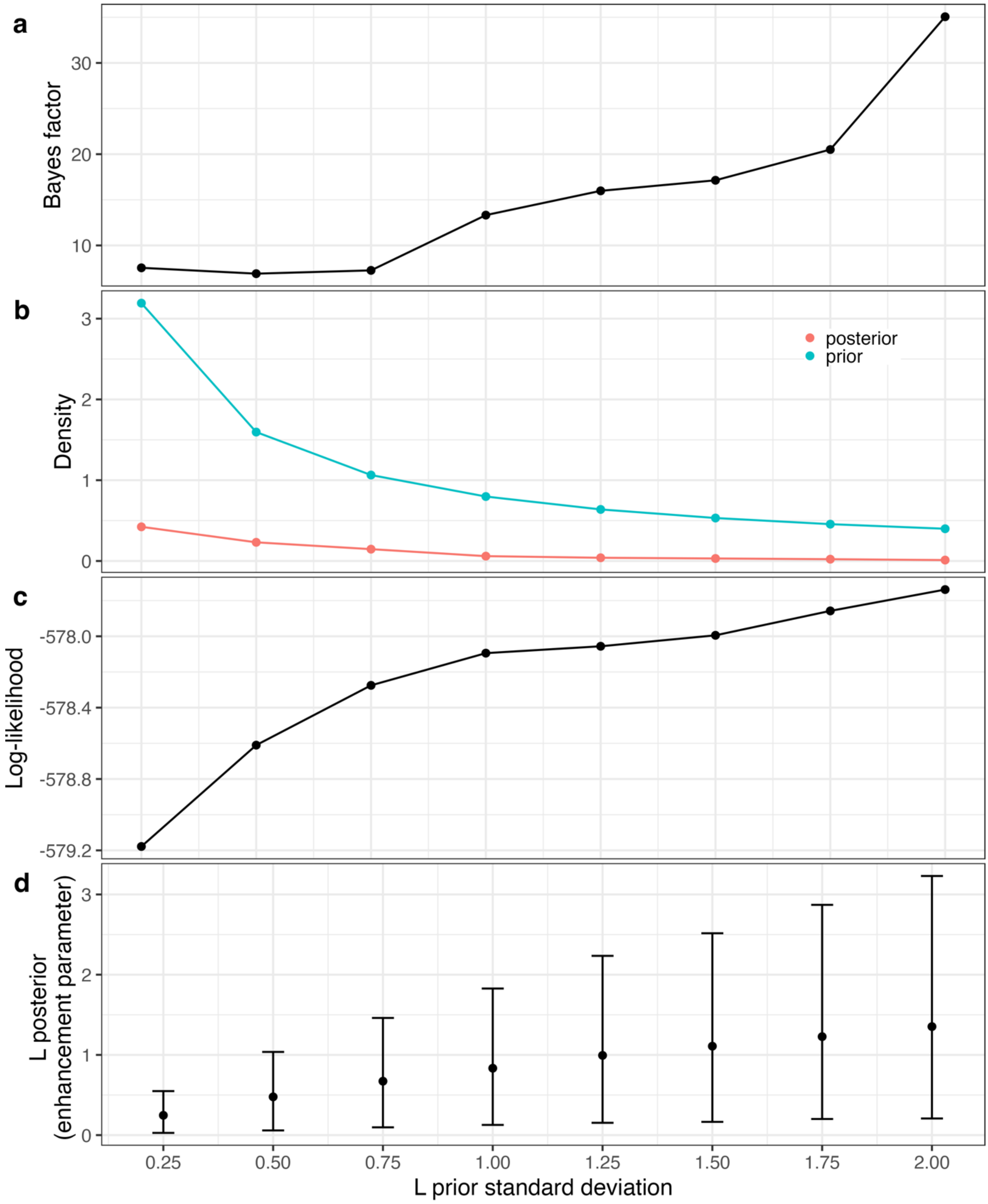

**Supplementary Figure 11: Sensitivity analysis on the impact of the choice of the prior distribution of the enhancement parameter (L) on the model support for vaccine-associated enhancement.** All prior distributions are truncated normal, centred on 0 (no enhancement). Standard deviations of the prior ranged from 0.25 to 2.00. **(a)** Bayes factor estimates comparing the vaccine efficacy model with and without enhancement (L fixed at 0). **(b)** Mean of the prior and posterior densities at 0. **(c)** Mean log-likelihood of the model with enhancement. **(d)** Posterior mean (point) and 95% credible interval (error bar) of the enhancement parameter L.

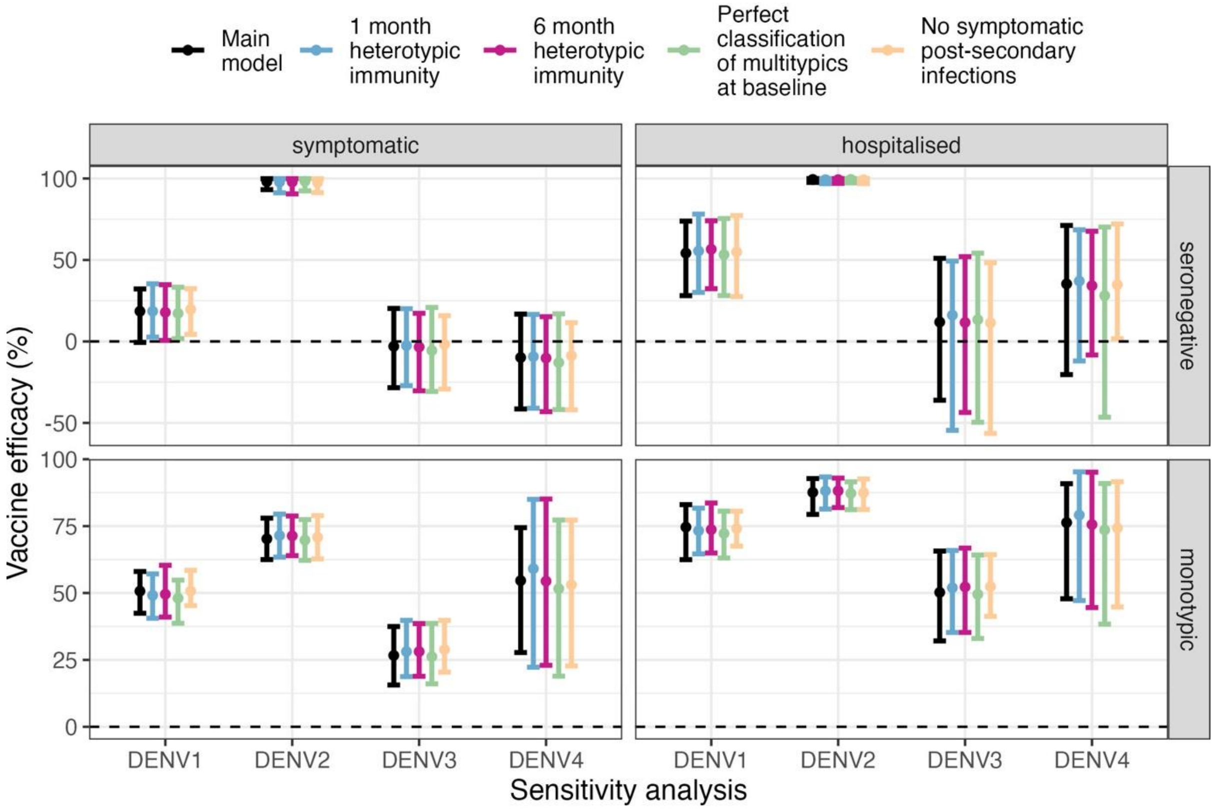

**Supplementary Figure 12**: **Sensitivity analysis**. Vaccine efficacy estimates for each sensitivity analysis (colours) by serotype (x-axis), serostatus (rows), against symptomatic disease and hospitalisation (columns). The main model assumes that heterotypic immunity lasts 12 months, multitypic individuals can be misclassified as seronegative at baseline, and post-secondary infections can be symptomatic. Point and error bars are respectively the mean and 95% credible interval of 100 posterior distribution samples. The dashed horizontal line marks 0 efficacy. Vaccine efficacy estimates in multitypic individuals are not plotted.

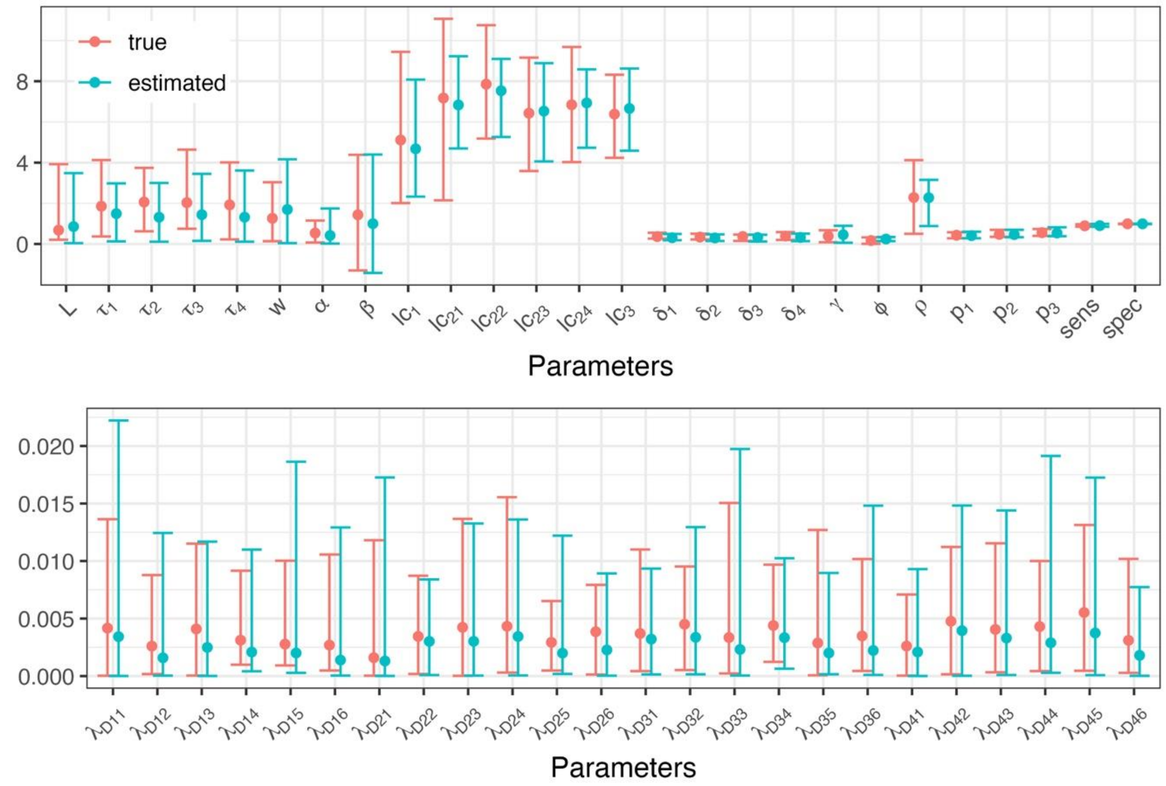

**Supplementary Figure 13**: **Model validation to simulated case data.** Pink point and error bars represent respectively the median and 95% confidence interval of 20 true parameter sets used to simulate the case data. Blue point and error bars are respectively the median and 95% credible interval of the posterior distribution obtained by fitting the model to the simulated case data. See **Supplementary Table 2** for a full description of the parameters used.

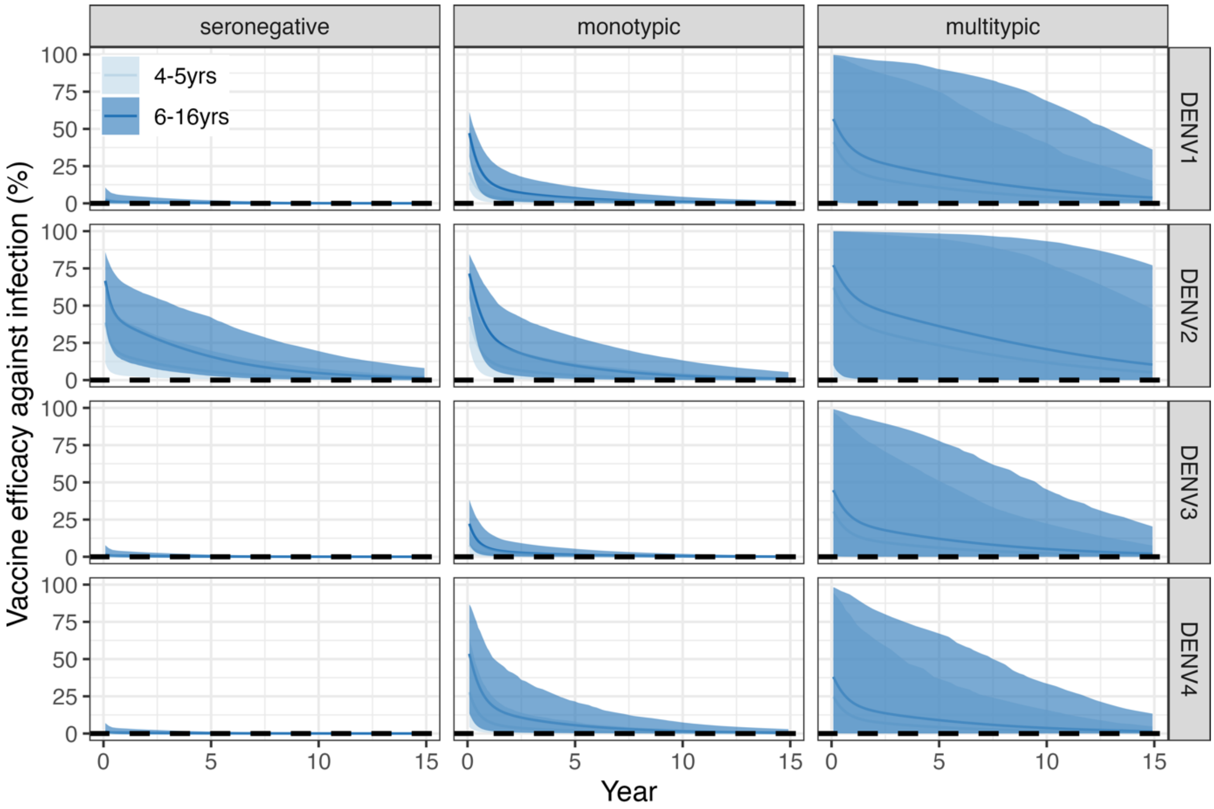

**Supplementary Figure 14: Vaccine efficacy against infection scenario.** Assumed VE against infection by age (colours), serotype (rows) and serostatus (columns). The solid line represents the mean and the shaded area represents the 95% CrI. The dashed horizontal line marks 0 efficacy.

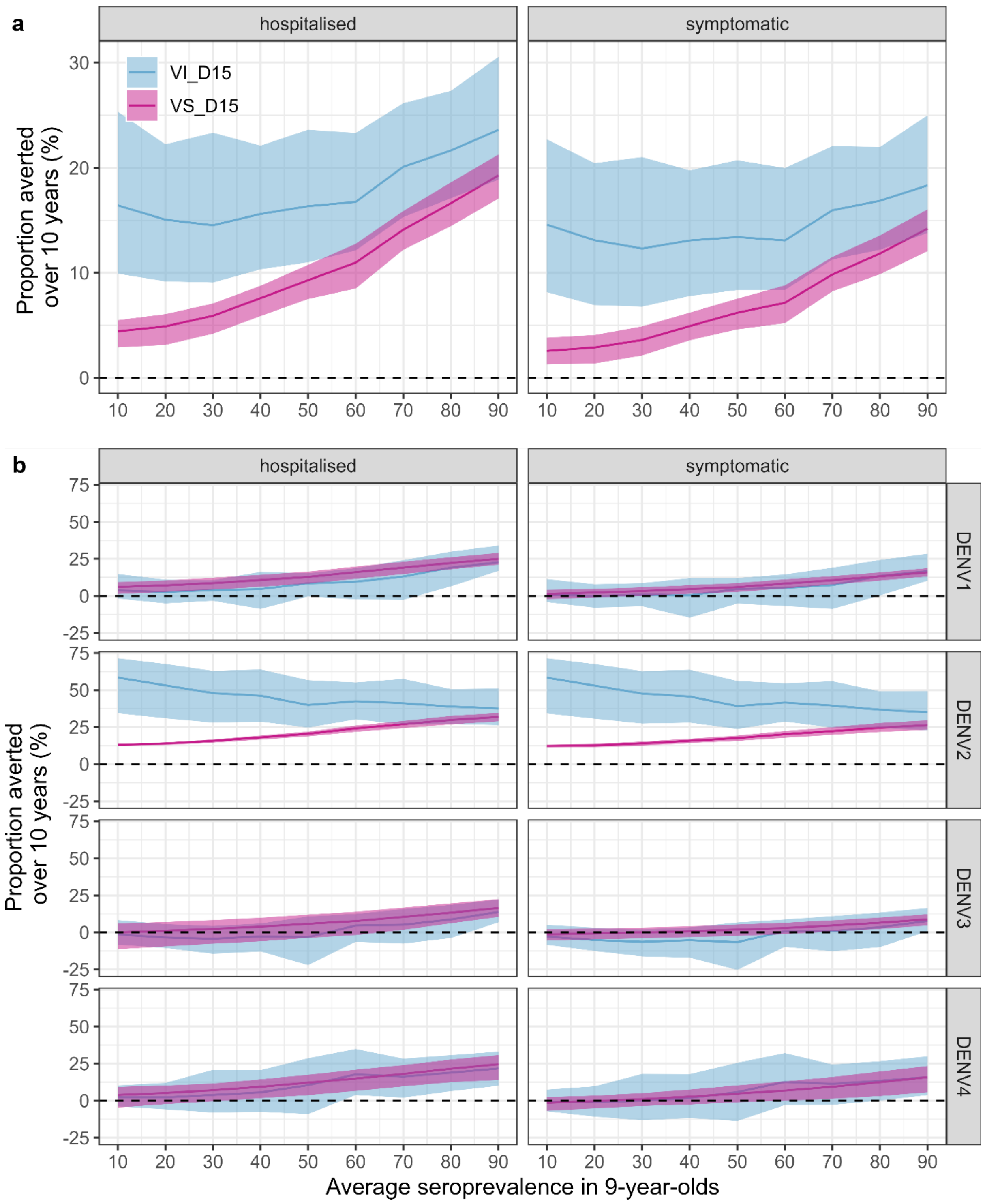

**Supplementary Figure 15: Population-level impact of vaccination in the Philippines.** Cumulative proportion of hospitalised and symptomatic cases averted (y-axis) by transmission setting (x-axis, expressed as the average seroprevalence in 9-year-olds), assuming efficacy against infection and disease (VI, blue) or only disease (VS, pink) decaying for 15 years (D15), using 80% coverage across ten years and the Philippines demography **(a)** over all serotypes and **(b)** by serotype. The solid line represents the mean, and the shaded regions represent the 95% credible interval.

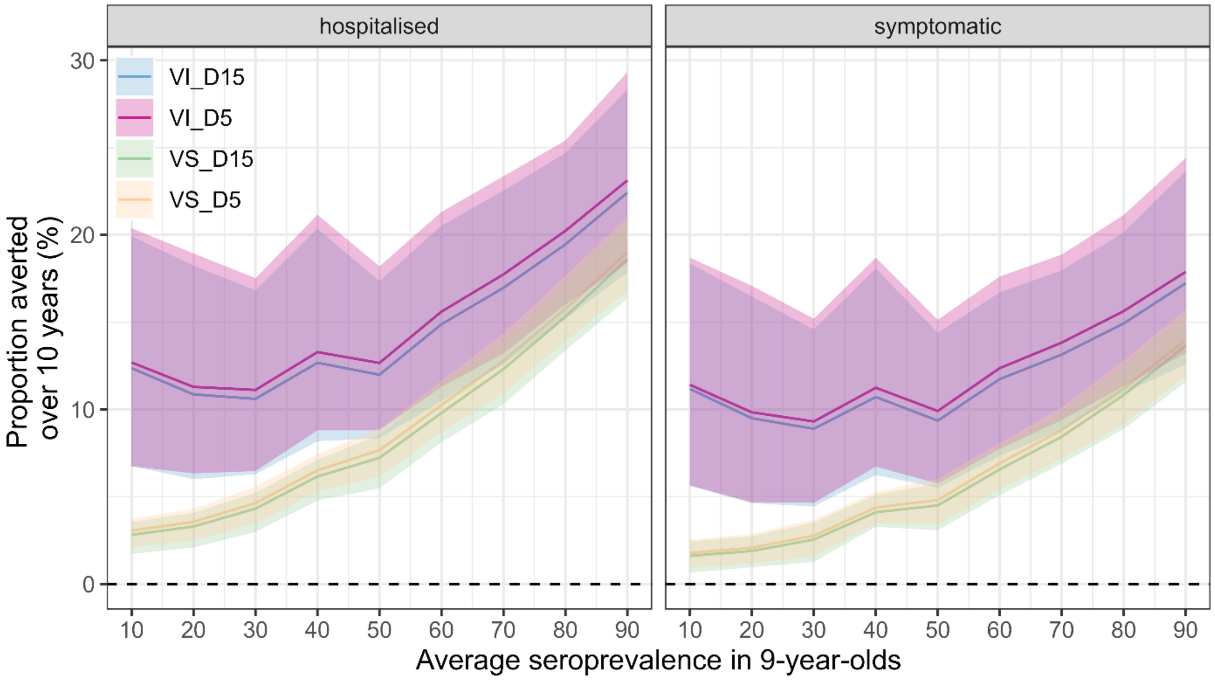

**Supplementary Figure 16: Population-level impact of vaccination in Brazil by the different scenarios of vaccine mechanism of action.** Cumulative proportion of hospitalised and symptomatic cases averted (y-axis) by transmission setting (x-axis, expressed as the average seroprevalence in 9-year-olds), assuming 80% coverage across ten years, assuming efficacy against infection and disease decaying for 5 years (VI_D5, pink), infection and disease decaying for 15 years (VI_D15, blue), only disease decaying for 5 years (VS_D5, yellow), only disease decaying for 15 years (VS_D15, green). The solid line represents the mean, and the shaded regions represent the 95% credible interval.

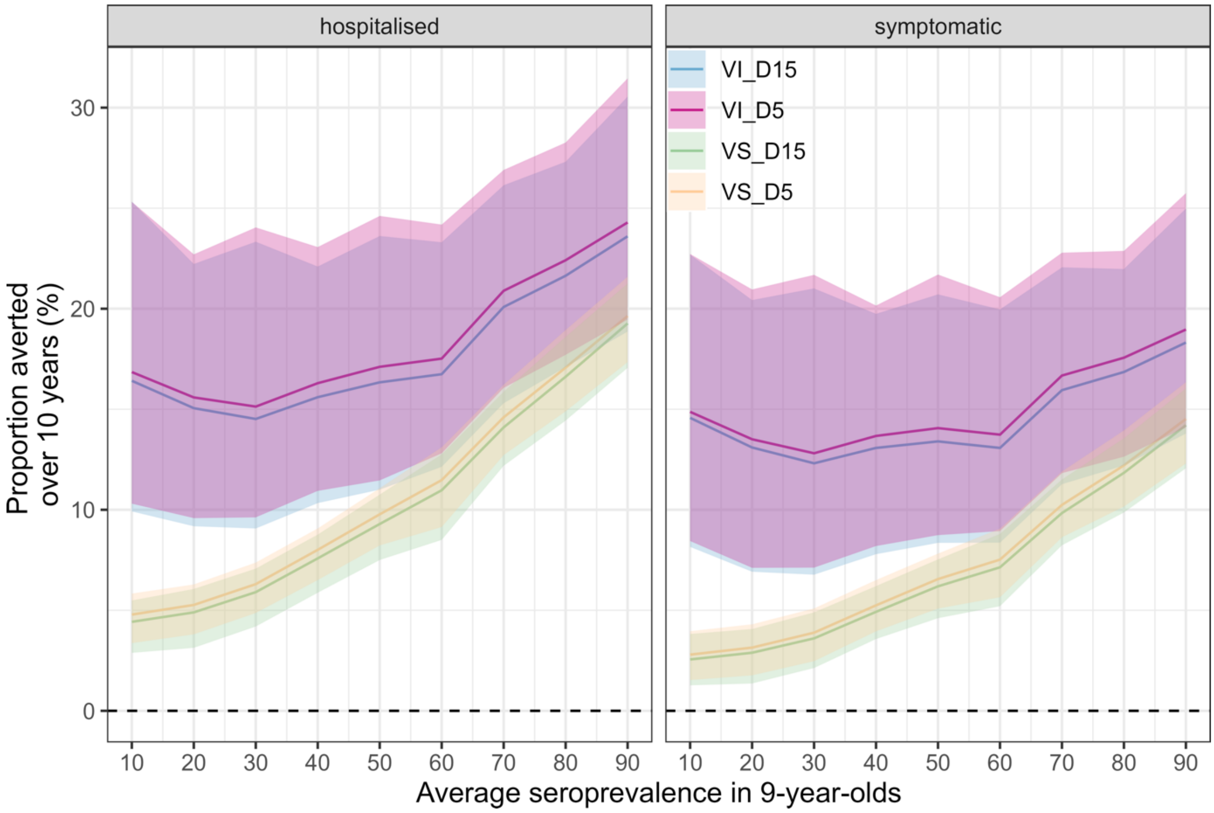

**Supplementary Figure 17: Population-level impact of vaccination in the Philippines by the different scenarios of vaccine mechanism of action.** Cumulative proportion of hospitalised and symptomatic cases averted (y-axis) by transmission setting (x-axis, expressed as the expected seroprevalence in 9-year-olds), assuming 80% coverage across ten years, assuming efficacy against infection and disease decaying for 5 years (VI_D5, pink), infection and disease decaying for 15 years (VI_D15, blue), only disease decaying for 5 years (VS_D5, yellow), only disease decaying for 15 years (VS_D15, green). The solid line represents the mean, and the shaded regions represent the 95% credible interval.

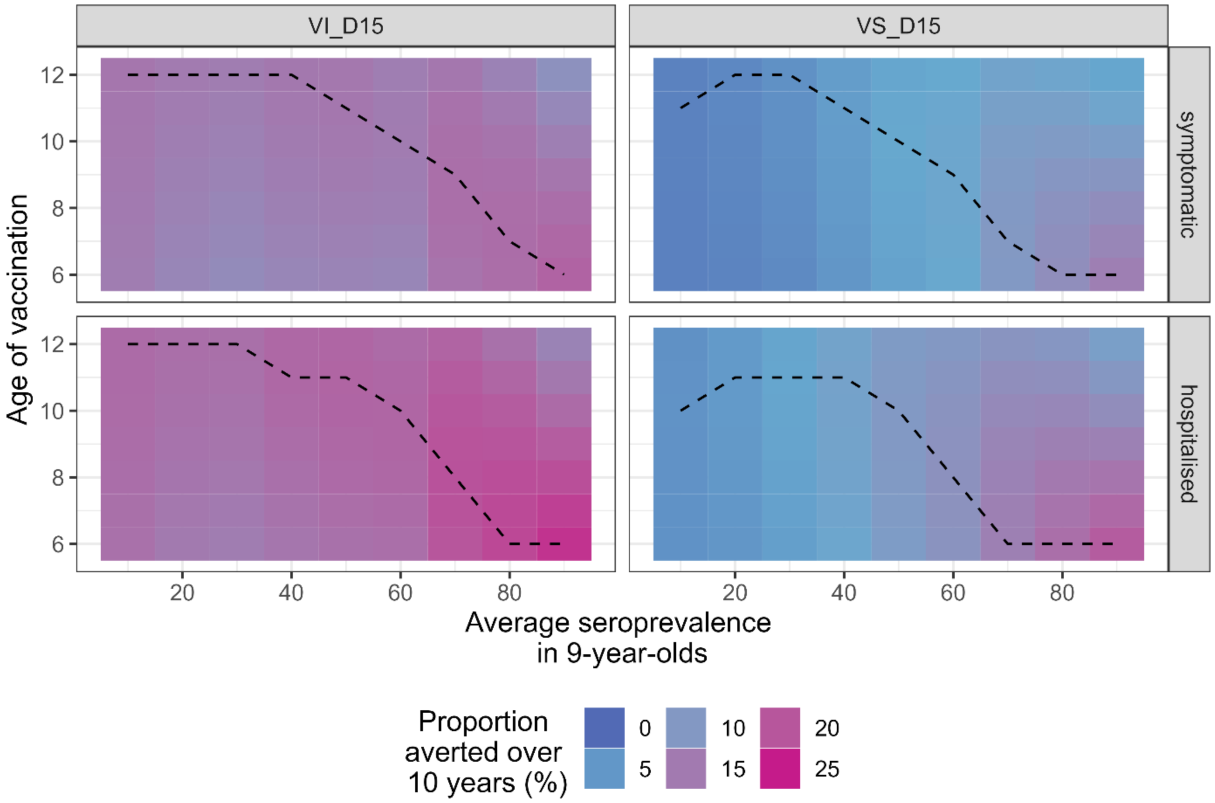

**Supplementary Figure 18: Impact of the age at vaccination on the population-level impact in the Philippines by transmission setting.** Cumulative proportion of hospitalised and symptomatic cases averted (rows) by transmission setting (x-axis, expressed as the average seroprevalence at 9-year-old), and vaccine mechanism (columns) assuming vaccination of ages 6-12 (y-axis) and the Philippines demography, over ten years. VI_D15: scenario assuming efficacy against infection and disease decaying for 15 years post vaccination. VS_D15: scenario assuming efficacy against disease decaying for 15 years post vaccination.

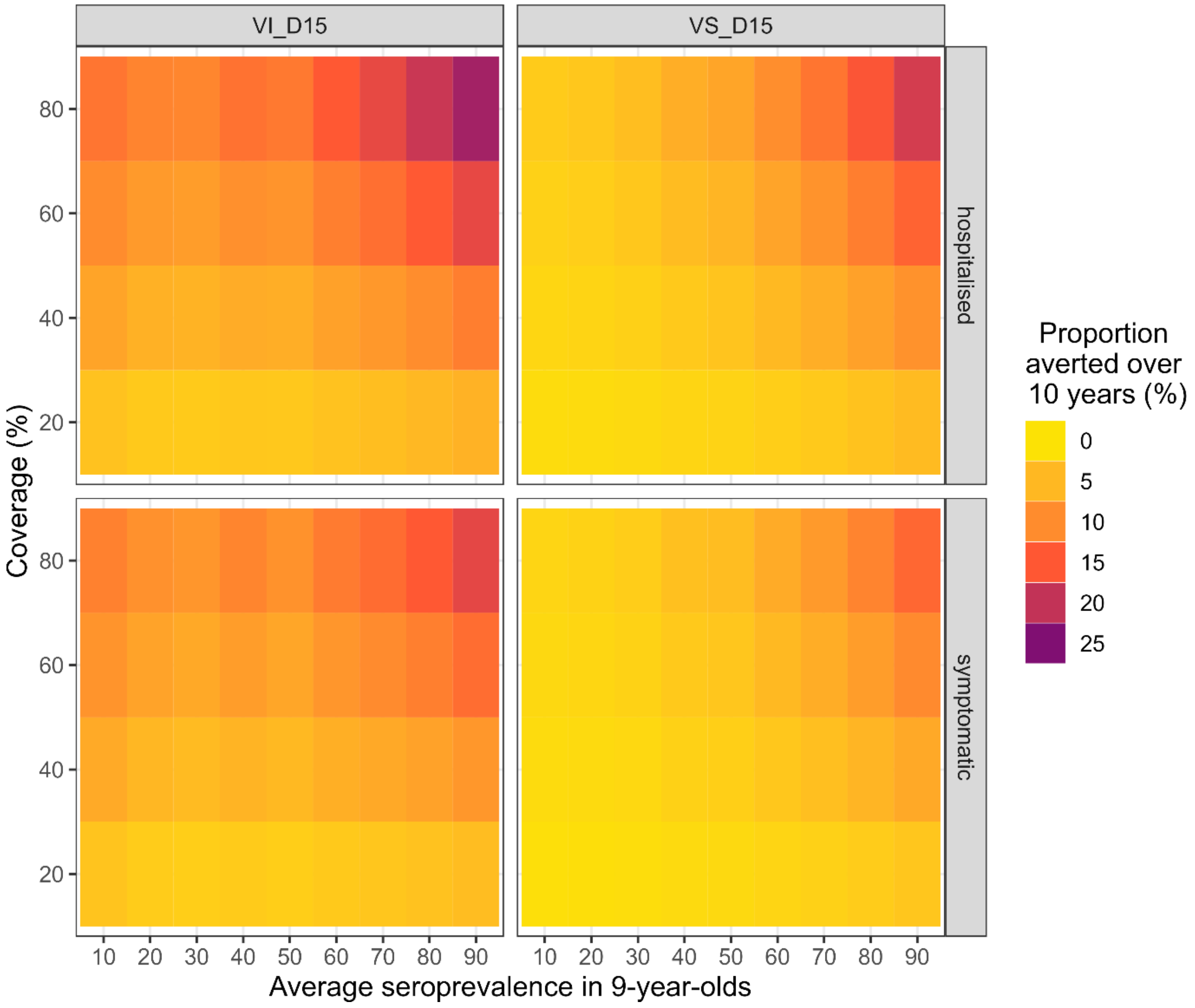

**Supplementary Figure 19: Effect of vaccination coverage on the population-level impact in Brazil.** Cumulative proportion of hospitalised and symptomatic cases averted (rows) by transmission setting (x-axis, expressed as the average seroprevalence at 9-year-old), and vaccine mechanism (columns) assuming 20-80% coverage (y-axis) and the Brazilian demography, over ten years.

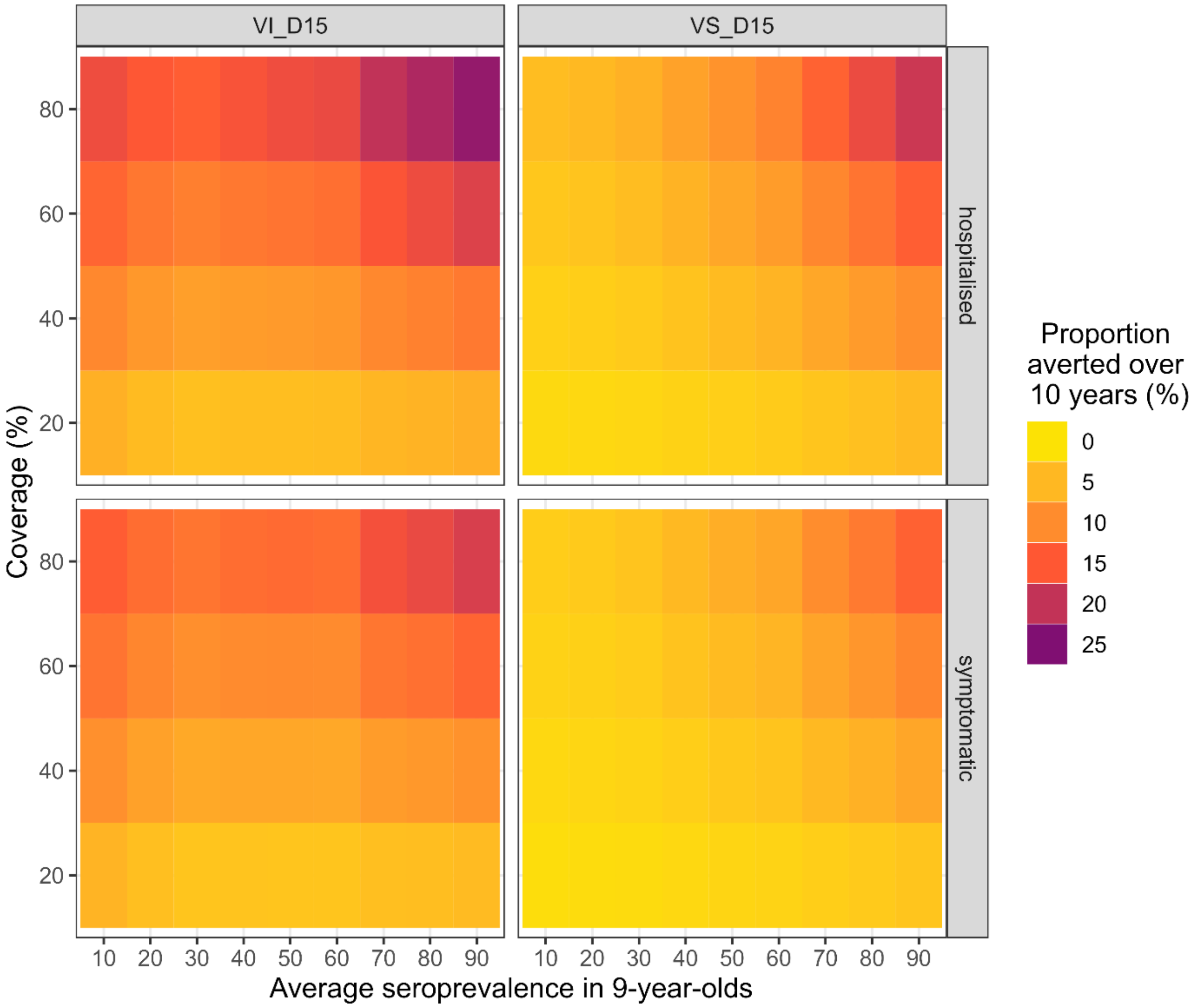

**Supplementary Figure 20: Effect of vaccination coverage on the population-level impact in the Philippines.** Cumulative proportion of hospitalised and symptomatic cases averted (rows) by transmission setting (x-axis, expressed as the average seroprevalence at 9-year-old), and vaccine mechanism (columns) assuming 20-80% coverage (y-axis) and the Philippines demography, over ten years.

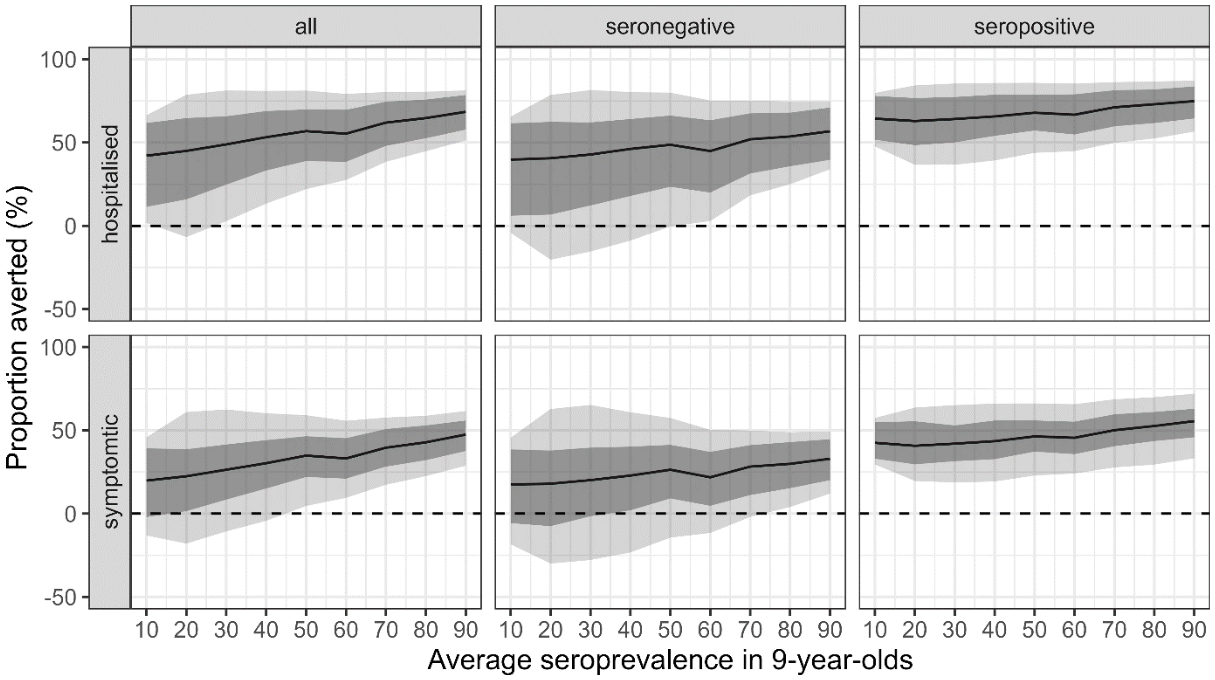

**Supplementary Figure 21: Individual-level impact of vaccination in the Philippines assuming efficacy against disease only (VS_D15).** Proportion of hospitalised and symptomatic cases averted (rows) in the first vaccinated cohort of 6-year-olds over ten years by transmission setting, expressed as the average seroprevalence at 9-year-old (x-axis) assuming a vaccination coverage of 80% using model VS_D15 and the Philippines demography. The impact is shown overall (all) and among baseline seropositive and seronegative vaccinees (columns). The solid lines represent the mean, light shading represents the overall uncertainty (95% CrI), and the dark shading represents the parameter uncertainty (95% CrI).

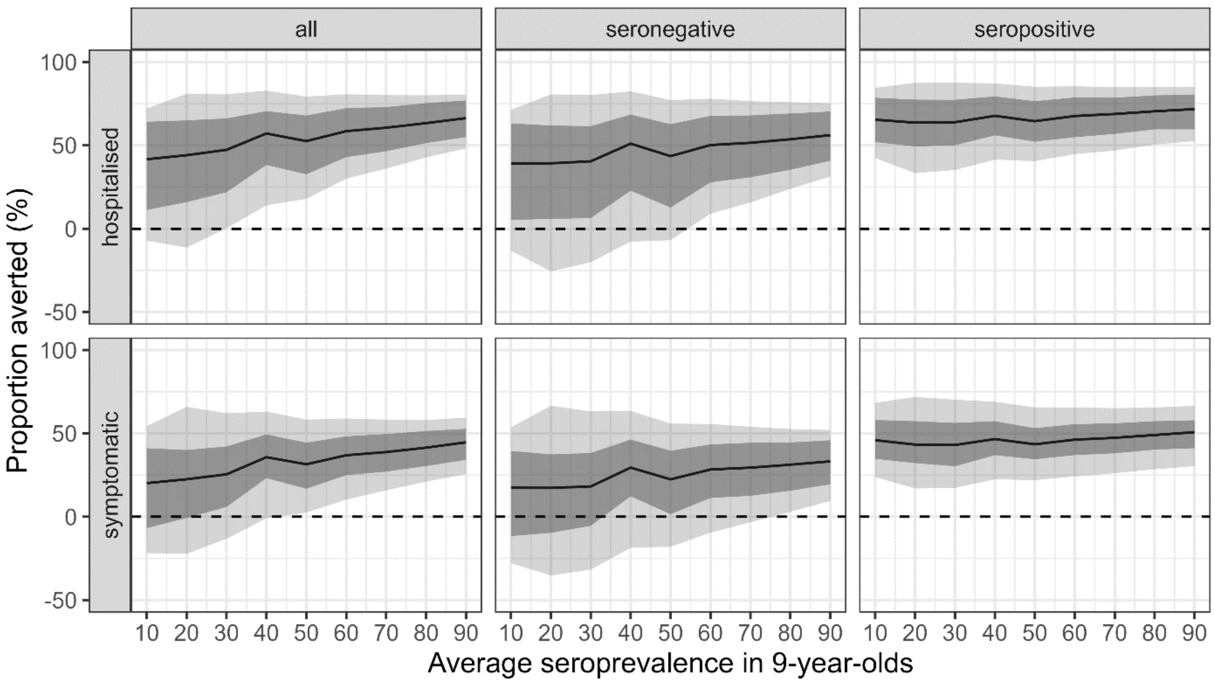

**Supplementary Figure 22: Individual-level impact of vaccination in Brazil assuming vaccine efficacy against infection and disease (VI_D15).** Proportion of hospitalised and symptomatic cases averted (rows) in the first vaccinated cohort of 6-year-olds over ten years by transmission setting, expressed as the average seroprevalence at 9-year-old (x-axis) assuming a vaccination coverage of 80% using model VI_D15 and the Brazil demography. The impact is shown overall (all) and among baseline seropositive and seronegative vaccinees (columns). The solid lines represent the mean, light shading represents the overall uncertainty (95% CrI), and the dark shading represents the parameter uncertainty (95% CrI).

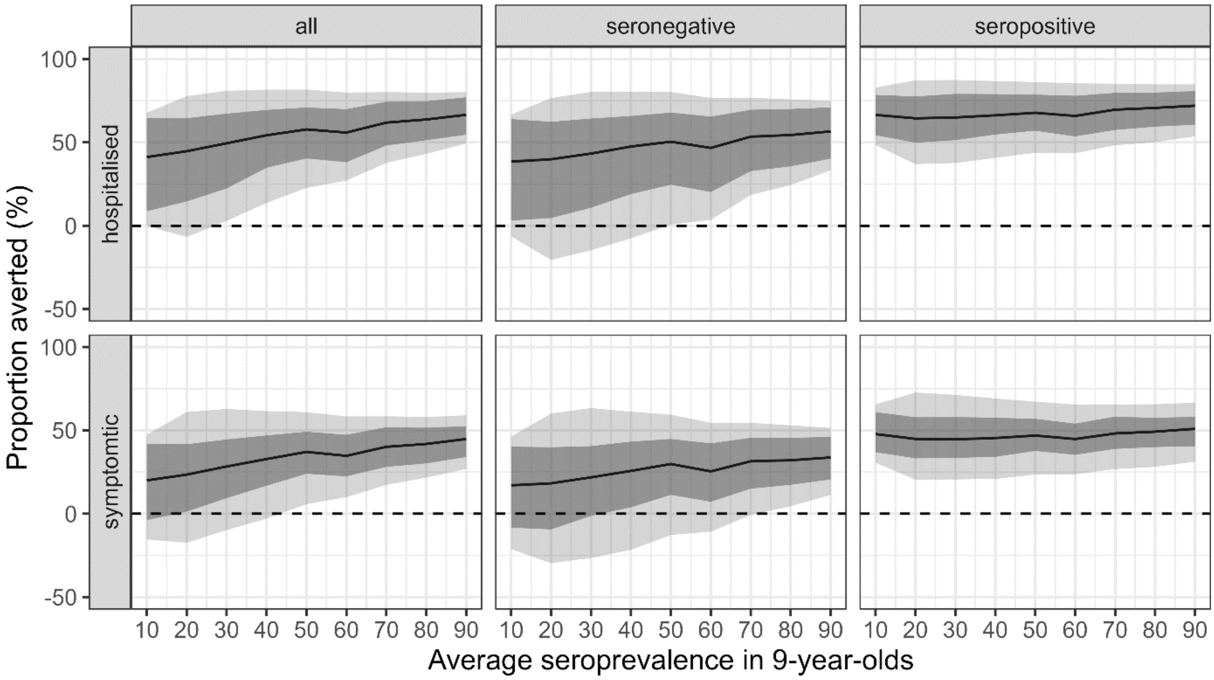

**Supplementary Figure 23: Individual-level impact of vaccination in the Philippines assuming vaccine efficacy against infection and disease (VI_D15).** Proportion of hospitalised and symptomatic cases averted (rows) in the first vaccinated cohort of 6-year-olds over ten years by transmission setting, expressed as the average seroprevalence at 9-year-old (x-axis) assuming a vaccination coverage of 80% using model VI_D15 and the Philippines demography. The impact is shown overall (all) and among baseline seropositive and seronegative vaccinees (columns). The solid lines represent the mean, light shading represents the overall uncertainty (95% CrI), and the dark shading represents the parameter uncertainty (95% CrI).

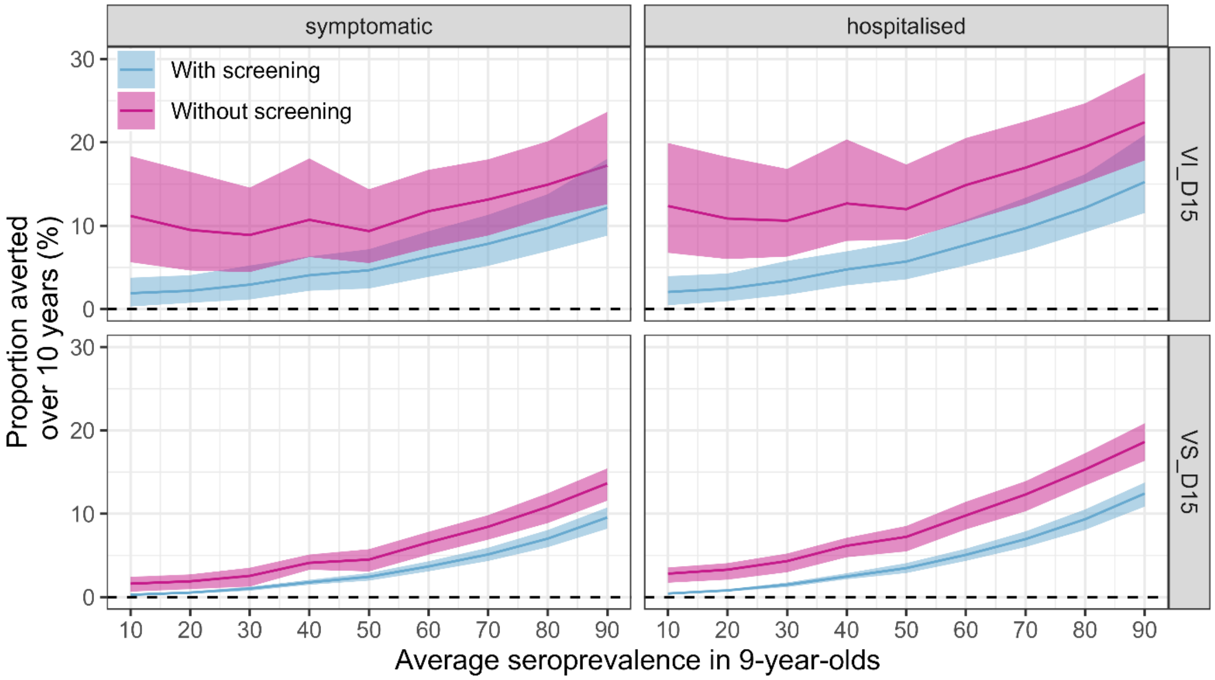

**Supplementary Figure 24: Population impact of pre-vaccination screening in Brazil.** Proportion of symptomatic cases and hospitalisations averted (y-axis) with (blue) and without (pink) pre-vaccination screening, over 10 years since the start of routine vaccination in the entire population by transmission setting (x-axis, expressed as the average 9-year-old seroprevalence), assuming 80% coverage and scenario VS_D15 or VI_D15 (rows).

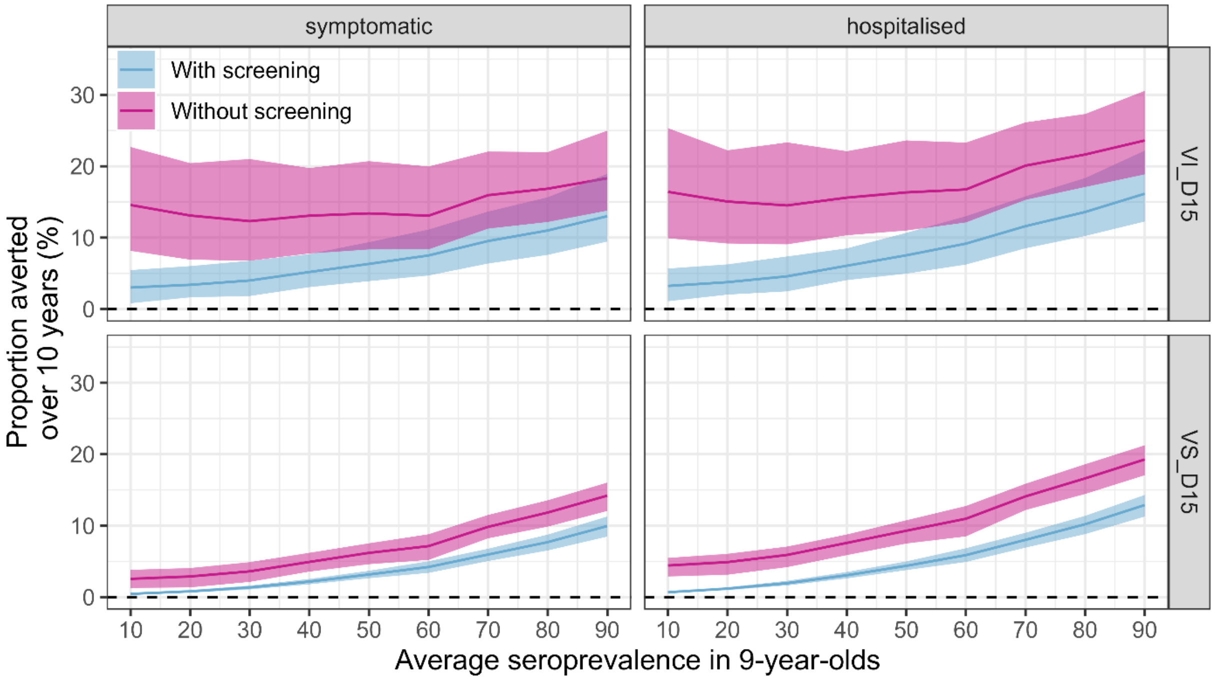

**Supplementary Figure 25: Population impact of pre-vaccination screening in the Philippines.** Proportion of symptomatic cases and hospitalisations averted (y-axis) with (blue) and without (pink) pre-vaccination screening, over 10 years since the start of routine vaccination in the entire population by transmission setting (x-axis, expressed as the average 9-year-old seroprevalence), assuming 80% coverage and scenario VS_D15 or VI_D15 (rows).

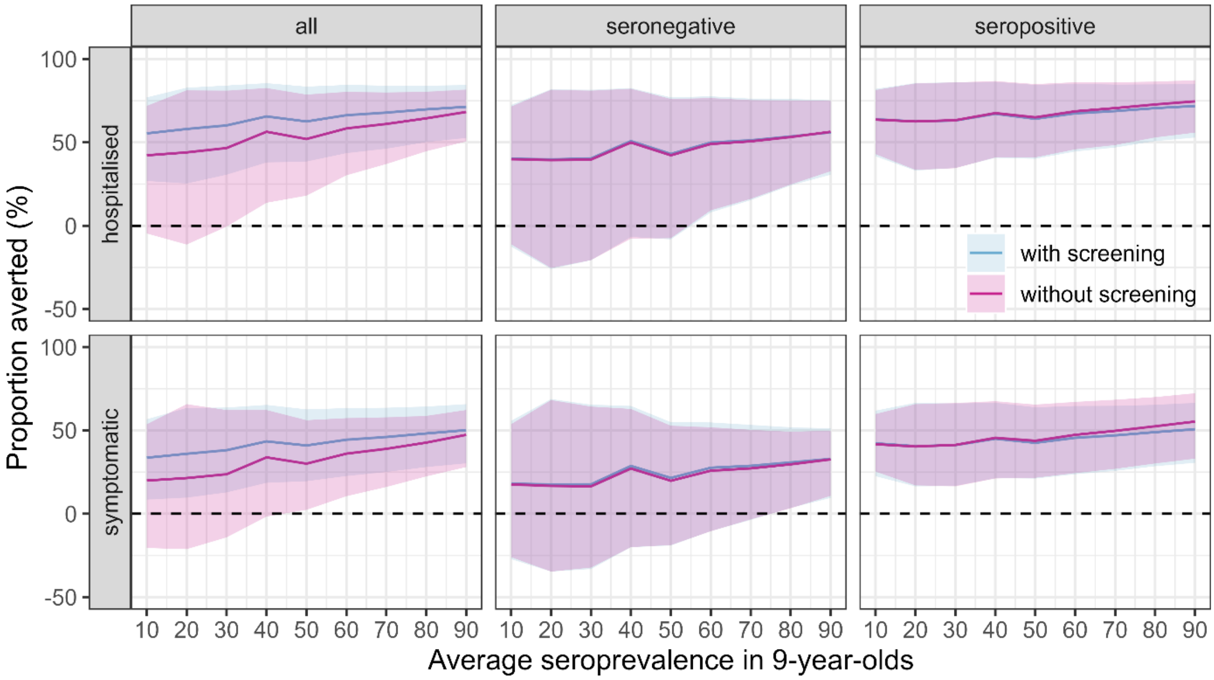

**Supplementary Figure 26: Individual-level impact of pre-vaccination screening in Brazil.** Proportion of hospitalised and symptomatic cases averted (rows) in the first vaccinated cohort of 6-year-olds over ten years by transmission setting, expressed as the average seroprevalence at 9-year-old (x-axis) assuming a vaccination coverage of 80% using model VS_D15 and the Brazil demography. The impact is shown overall (all) and among baseline seropositive and seronegative vaccinees (columns). The solid lines represent the mean and shaded region represents the overall uncertainty (95% CrI).

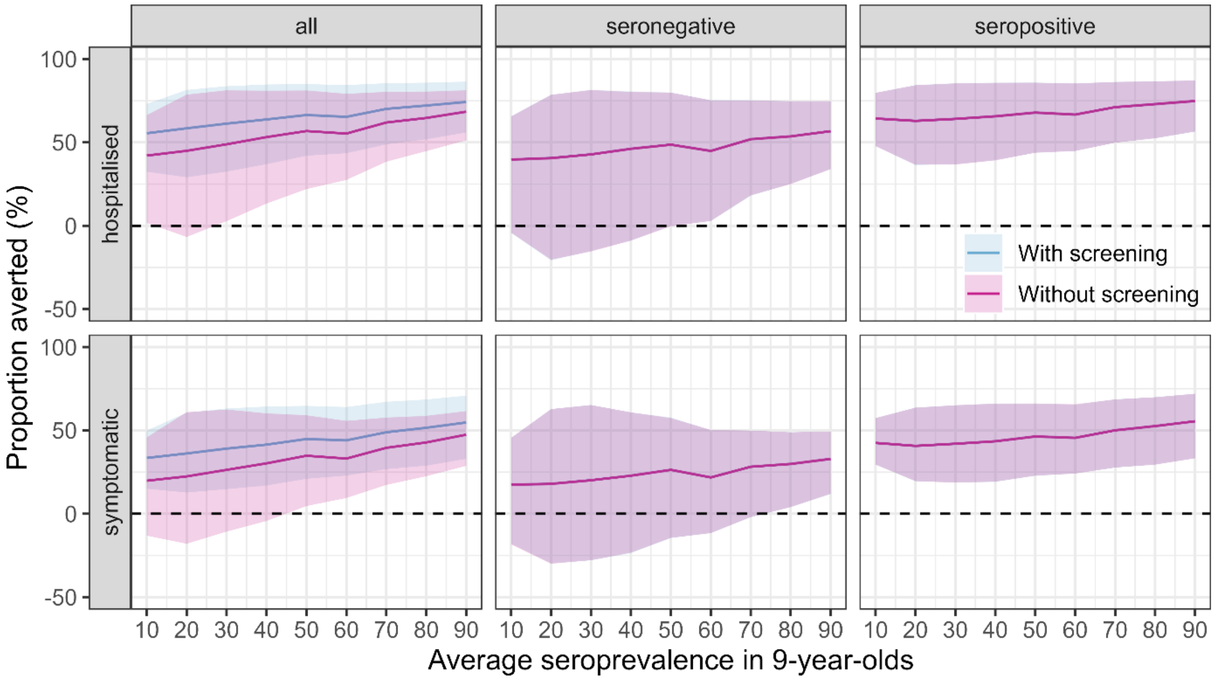

**Supplementary Figure 27: Individual-level impact of pre-vaccination screening in the Philippines.** Proportion of hospitalised and symptomatic cases averted (rows) in the first vaccinated cohort of 6-year-olds over ten years by transmission setting, expressed as the average seroprevalence at 9-year-old (x-axis) assuming a vaccination coverage of 80% using model VS_D15 and the Philippines demography. The impact is shown overall (all) and among baseline seropositive and seronegative vaccinees (columns). The solid lines represent the mean and shaded region represents the overall uncertainty (95% CrI).

#### References

1. United Nations. World Population Prospects. (2022).
2. Medina, F. A. et al. Comparison of the Sensitivity and Specificity of Commercial Anti-Dengue Virus IgG Tests to Identify Persons Eligible for Dengue Vaccination. 2024.04.19.24306097 Preprint at https://doi.org/10.1101/2024.04.19.24306097 (2024).
3. El Hindi, T. et al. Estimated efficacy of TAK-003 against asymptomatic dengue infection in children and adolescents. (2023).
4. Salje, H. et al. Evaluation of extended efficacy of Dengvaxia vaccine against symptomatic and subclinical dengue infection. Nat Med 27, 1395–1400 (2021).
5. Salje, H. et al. Reconstruction of antibody dynamics and infection histories to evaluate dengue risk. Nature 557, 719–723 (2018).
6. Ferguson, N. M. et al. Benefits and risks of the Sanofi-Pasteur dengue vaccine: Modeling optimal deployment. Science 353, 1033–1036 (2016).
